# Large Registry Based Analysis of Genetic Predisposition to Tuberculosis Identifies Genetic Risk Factors at HLA

**DOI:** 10.1101/2022.01.27.22269951

**Authors:** Anniina Tervi, Nella Junna, Martin Broberg, Samuel E. Jones, FinnGen, Satu Strausz, Hanna-Riikka Kreivi, Caroline A. Heckman, Hanna M. Ollila

## Abstract

Tuberculosis is a significant public health concern resulting in the death of over 1 million individuals each year worldwide. While treatment options and vaccines exist, a substantial number of infections still remain untreated or are caused by treatment resistant strains. Therefore, it is important to identify mechanisms that contribute to risk and prognosis of tuberculosis as this may provide tools to understand disease mechanisms and provide novel treatment options for those with severe infection. Our goal was to identify genetic risk factors that contribute to the risk of tuberculosis and to understand biological mechanisms and causality behind the risk of tuberculosis. A total of 1,895 individuals in the FinnGen study had ICD-based tuberculosis diagnosis. GWAS analysis identified genetic variants with statistically significant association with tuberculosis at the Human leukocyte antigen (HLA) region (p<5e-8) and at rs560595454 in gene *INPP5A*. Fine mapping the HLA-association provided evidence for one protective haplotype tagged by *HLA DQB1*05:01* (p=1.82E-06, OR = 0.81 [CI 95 % 0.74-0.88]), and predisposing alleles tagged by *HLA DRB1*13:02* (p=0.00011, OR = 1.35 [CI 95% 1.16-1.57]). Furthermore, genetic correlation analysis showed association with earlier reported risk factors including smoking (p<0.05). Mendelian randomization supported smoking as a risk factor for tuberculosis (inverse-variance weighted p<0.05, OR = 1.83 [CI 95 % 1.15-2.93]) with no significant evidence of pleiotropy. Our findings indicate that specific HLA alleles and *INPP5A* associate with the risk of tuberculosis. In addition, lifestyle risk factors such as smoking contribute to the risk of developing tuberculosis.

## Introduction

Tuberculosis (TB) is an infectious disease caused by *Mycobacterium tuberculosis*. The bacteria are transmitted via airborne transmission mainly affecting the pulmonary organs (pulmonary TB) but they can also affect other organs (extra-pulmonary TB) (Pai *et al*., 2016; Global Tuberculosis Report, 2020). TB can manifest as a latent non-transmissible form or as an active transmissible form where patients experience symptoms such as high fever, persistent cough, fatigue and weight loss (Pai *et al*., 2016). Most individuals infected by *Mycobacterium tuberculosis* will not develop TB whereas 5 to 10 % of infected individuals develop the disease. Individuals with a compromised immune system, such as those infected with HIV (human immunodeficiency virus) have a higher likelihood of developing TB (Global Tuberculosis Report, 2020).

Incidence rates of TB in most of Europe and Northern America are less than 10 per 100,000 per year, and mortality in HIV-negative people is less than 5 per 100 000 per year (Global Tuberculosis Report, 2020). After development of BCG (Bacillus Calmette–Guérin) vaccination in 1921, people were widely vaccinated in Europe, which lead to declining TB incidence rates. Decline was further assisted by development of antibiotic treatments in the 1940’s and 1950’s, after which latent TB infections could be treated (Daniel, 2006; Langer *et al*. 2019). However, Europe has the highest rate of new reported cases of multidrug resistant TB (MDR-TB) (Global Tuberculosis Report, 2020).

The risk for TB is also highly correlated with comorbid diseases. While HIV infection is the strongest risk factor for TB disease, TB infection also exacerbates the course of HIV infection and is associated with 4-fold all-cause mortality among HIV-patients (Bell and Noursadeghi, 2018). Similarly, malnutrition as well as deficiency of vitamins C or D have been associated with TB disease (Cegielski and McMurray, 2004; Sinha *et al*., 2019) although this association is also related with overall fragility: very young and very old people have elevated TB risk (reviewed in Marais *et al*., 2013). Finally, smoking increases risk for latent TB infection and TB disease more than 2-fold (Bates *et al*., 2007), and public health campaigns against smoking have been applied in high risk areas for TB.

On a global scale, WHO estimated that 10 million people developed TB disease in the year 2019 and from those estimated 1.2 million HIV-negative individuals died due to TB in 2019 (Global Tuberculosis Report, 2020). Although there are treatments for TB, long term consequences such as pulmonary dysfunction and prolonged respiratory symptoms may expose survivors to other respiratory disorders, for instance, to chronic obstructive pulmonary disease (COPD) (Lam *et al*., 2010; Akkara *et al*., 2013; Ralph *et al*., 2013; Nihues *et al*., 2015). In addition, treatment resistant strains, comorbidity burden create challenges in managing TB at the local and global scale.

Therefore, it is crucial to understand the biological mechanisms that contribute to onset of TB. Understanding these mechanisms is likely to result in better disease management and potentially novel treatment options as well as non-pharmaceutical measures to prevent and to treat TB infections.

By using genetic tools, we are able to study the biological mechanisms underlying TB along with causal relationships between TB and different comorbidities. Host genetic factors in TB have been studied in various different populations (for example Amirzargar *et al*., 2004; Kim *et al*., 2005; Thye *et al*., 2012; Curtis *et al*., 2015; Sveinbjornsson *et al*., 2016; Tian et al., 2017; Sobota *et al*., 2017; Qi *et al*., 2017; Quistrebert *et al*., 2021; Li *et al*., 2021). In European ancestry *Mycobacterium tuberculosis* or pulmonary TB GWAS (Sveinbjornsson *et al*., 2016) three variants rs557011, rs9271378 and rs9272785 in the HLA-region reached genome-wide significance (p<5E-08). Additionally, in self-reported positive TB test result GWAS, variant rs2894257 (also from the HLA-region) was found to be genome-wide significant in European ancestry individuals (Tian et al., 2017).

In this study, we used data from the FinnGen study data freeze 7 (R7) to explore the biological mechanisms underlying TB in 310,000 individuals. We were especially interested in assessing host genetic components using genome-wide association study (GWAS), and exploring comorbidity burden using genetic correlations, epidemiological tools, and causality with Mendelian randomization (MR).

## Materials and Methods

### Study Cohort

The FinnGen study (https://www.finngen.fi/en) is a public-private partnership including Finnish universities, biobanks and hospital districts together with several pharmaceutical companies founded in the year 2017. The aim is to collect both National Health Record and genetic data from 500,000 Finns. The study participants include patients with acute and chronic diseases as well as healthy voluntary and population collections. R7 includes ∼310,000 individuals (approximately 175,000 females and 135,000 males).

### FinnGen Ethics Statement

Patients and control subjects in FinnGen provided informed consent for biobank research, based on the Finnish Biobank Act. Alternatively, separate research cohorts, collected prior the Finnish Biobank Act came into effect (in September 2013) and start of FinnGen (August 2017), were collected based on study-specific consents and later transferred to the Finnish biobanks after approval by Fimea (Finnish Medicines Agency), the National Supervisory Authority for Welfare and Health. Recruitment protocols followed the biobank protocols approved by Fimea. The Coordinating Ethics Committee of the Hospital District of Helsinki and Uusimaa (HUS) statement number for the FinnGen study is Nr HUS/990/2017.

The FinnGen study is approved by Finnish Institute for Health and Welfare (permit numbers: THL/2031/6.02.00/2017, THL/1101/5.05.00/2017, THL/341/6.02.00/2018, THL/2222/6.02.00/2018,THL/283/6.02.00/2019, THL/1721/5.05.00/2019 and THL/1524/5.05.00/2020), Digital and population data service agency (permit numbers: VRK43431/2017-3, VRK/6909/2018-3, VRK/4415/2019-3), the Social Insurance Institution (permit numbers: KELA 58/522/2017, KELA 131/522/2018,KELA 70/522/2019, KELA 98/522/2019, KELA 134/522/2019, KELA 138/522/2019, KELA 2/522/2020,KELA 16/522/2020), Findata permit numbers THL/2364/14.02/2020, THL/4055/14.06.00/2020, THL/3433/14.06.00/2020, THL/4432/14.06/2020, THL/5189/14.06/2020, THL/5894/14.06.00/2020, THL/6619/14.06.00/2020, THL/209/14.06.00/2021, THL/688/14.06.00/2021, THL/1284/14.06.00/2021,THL/1965/14.06.00/2021, THL/5546/14.02.00/2020 and Statistics Finland (permit numbers: TK-53-1041-17 and TK/143/07.03.00/2020 (earlier TK-53-90-20)).

The Biobank Access Decisions for FinnGen samples and data utilized in FinnGen Data Freeze7 include: THL Biobank BB2017_55, BB2017_111, BB2018_19, BB_2018_34, BB_2018_67,

BB2018_71, BB2019_7, BB2019_8, BB2019_26, BB2020_1, Finnish Red Cross Blood Service Biobank 7.12.2017, Helsinki Biobank HUS/359/2017, Auria Biobank AB17-5154 and amendment #1 (August 17 2020), Biobank Borealis of Northern Finland_2017_1013, Biobank of Eastern Finland 1186/2018 and amendment 22 § /2020, Finnish Clinical Biobank Tampere MH0004 and amendments (21.02.2020 06.10.2020), Central Finland Biobank 1-2017, and Terveystalo Biobank STB 2018001.2.

### Phenotype Definition

In the FinnGen study, the main phenotype used in our study was TB of all organs defined using ICD-10 based diagnosis codes A15-A19. Individuals defined as a case in TB of all organs endpoint had to have at least one of the following ICD-10 codes or their subcode: A15, A16, A17, A18 or A19. Other phenotypes used were respiratory TB and TB of other organs. An individual was defined as a case in respiratory TB when that person had at least one of the following ICD-10 codes or their subcode: A15 or A16. An individual was defined as a case in TB of other organs when that person had the ICD-code A18 or one of its subcodes. All individuals in the cohort were Finns and matched against the SiSu v3 reference panel (http://www.sisuproject.fi/).

### HLA Imputation

HLA imputation was performed for *HLA A, HLA B, HLA C, HLA DRB1, HLA DQA1, HLA DQB1, HLA DPB1, HLA DRB3, HLA DRB4* and *HLA DRB5* using HIBAG, as implemented earlier (Ritari *et al*., 2020).

### Statistical Tools

For the FinnGen cohort, GWAS was conducted using the REGENIE pipeline (https://github.com/FINNGEN/regenie-pipelines). Manhattan-plot for FinnGen R7 GWASs were plotted using R version 4.0.1 (packages: qqman and RColorBrewer).

Association testing between individual HLA alleles and TB was conducted with multivariate logistic regression using R (version 4.0.3, packages: data.table, dplyr and tidyverse). Multivariate logistic regression model was adjusted for age at death or end of follow up (12/31/2019), sex and the first 20 genetic principal components (adjusting for principal components accounts for population structure within the cohort). Step-wise logistic regression was conducted by adding the most strongly associated HLA allele as a covariate to the multivariate regression analysis. This was repeated as many times as there were significant alleles left in the analysis.

The assessment of genetic correlation between TB and different traits, we performed linkage disequilibrium (LD) score regression analyses with LD HUB provided by the Broad Institute of MIT and Harvard and MRC Integrative Epidemiology Unit, University of Bristol (Zheng *et al*., 2017; Bulik-Sullivan *et al*., 2015a; Bulik-Sullivan *et al*., 2015b). Additionally, we tested alcohol use disorder (AUD) using LD score regression separately since the trait was not available in the LD HUB tool (Bulik-Sullivan *et al*., 2015a). For the LD score regression, we used the HapMap 3 SNP list and European LD score files provided with the software. Summary statistics for LD score regression were obtained from FinnGen R7 GWAS of TB of all organs and from Sanchez-Roige *et al*. (2019) for AUD.

We obtained the lead SNPs associated with smoking and used as exposure instruments against the FinnGen R7 TB GWAS summary statistics as ‘Cigarettes per day’ and ‘Age of initiation of regular smoking’ from a recent large-scale GWAS (Liu *et al*., 2019). The lead SNPs associated with vitamin D deficiency were obtained from the study by (Revez *et al*., 2020). The BMI associated SNPs and COPD associated SNPs were obtained from the IEU open GWAS project ID: ukb-b-19953. AUD associated SNPs were obtained from the study by (Sanchez-Roige *et al*., 2019). The MR was performed using the TwoSampleMR R package (Hemani *et al*., 2018; Hemani, Tilling, and Davey Smith, 2017). Furthermore, we tested for potential pleiotropic effects using the Egger intercept methods as part of the TwoSampleMR package and the MR-PRESSO package (Verbanck *et al*., 2018).

### Epidemiological Measures

Association testing between selected known risk factors and tuberculosis was conducted with multivariate logistic regression using R (version 4.0.3, packages: data.table, dplyr and tidyverse). Multivariate logistic regression model was adjusted with age at death or end of follow up (12/31/2019), BMI, sex and the first 10 genetic principal components. Phenotypic traits used: Smoking (ever vs. never), Smoking (current vs. former/never), Diabetes (ICD-10 codes E10-E14 or ATC-code (medicine purchases) A10B), Chronic obstructive pulmonary disease (COPD, ICD10-code J44), Hypertension (ICD10-codes I10-I15 or I67.4, or Finnish Social Insurance Institution (KELA) code 205), Asthma (ICD10-code J45), Major coronary heart disease event (CHD, ICD10-codes I20.0, I21 or I22), Human immunodeficiency virus (HIV, ICD10-codes B20-B24), Rheumatoid arthritis (RA, ICD10-codes M05 or M06), Biological medication for rheumatoid arthritis (ATC-codes: L04AA24, L04AC03, L04AB04, L04AB01, L04AB06, L04AB02, L04AB05, L04AA26, L01XC02, L04AC10, L04AC07 or L04AC05), Sleep apnea (ICD10-code G47.3), Alcohol dependence (ICD10-code F10.2), Alcohol use disorder (AUD, ICD10-codes F10.1 or F10.2), Inflammatory bowel disease (IBD, ICD10-codes K50, K51 or KELA codes 208 or 209) and Crohn’s disease (ICD10-code K50). Other known comorbidities such as vitamin D deficiency, organ transplantation and immunodeficiency were not included in the analyses due to the small number of cases in the FinnGen study.

A Kaplan-Meier estimator (Goel, Khanna, and Kishore, 2010) was used to create survival curves representing effect of selected comorbidity to survival among TB patients. A Cox proportional hazards model (Cox regression) was used to estimate the effect of selected risk factors among TB patients on survival (Cox, 1972). Cox regression was adjusted with stratified sex, stratified cohort (cohort representing for example biobank or study included within the FinnGen study), BMI and the first 10 genetic principal components. Survival function for the Kaplan-Meier estimator and Cox regression was constructed using age at death or end of follow up (12/31/2019) as time variable and death (0 or 1) as event variable. Additionally, proportional hazards assumption of Cox regression model was tested (Grambsch and Therneau, 1994). Analyses were conducted using R version 4.0.3 (packages: survival, survminer, survMisc, ggsurvplot and ggplot2).

## Results

### Genome-wide Association Study

To study the host genetic components contributing to TB we performed GWAS in 1,895 individuals with ICD-based TB and 307,259 controls from FinnGen R7 (Table 1, Figure 1). We identified four genome-wide significant (p<5E-08) single nucleotide polymorphisms (SNPs) of which two are at the HLA locus in the chromosomal position 6p21 (rs9391858 and rs33915496) and two are located in chromosome 10 (rs560595454 and rs562763274) in gene *INPP5A* (Inositol Polyphosphate-5-Phosphatase A) (Figure 2). The lead SNP (rs9391858) was closest to and downstream from gene *C6orf10* (open reading frame on chromosome 6, also known as *TSBP1* (testis expressed basic protein 1)), which is located in the HLA locus and upstream from *HLA DRA* gene (Figure S1-S2). We also performed GWAS on subsets of TB endpoints in FinnGen, respiratory TB (1,380 cases and 307,312 controls) and TB of other organs (538 cases and 307,259 controls), where a genome-wide significant SNP was identified in respiratory TB GWAS in the HLA locus in between *HLA DRA* and *HLA DQA1* genes (rs2395516, p=4.5E-09) (Figure S3-S4).

**Table 1.** FinnGen R7 Metrics for Tuberculosis.

|  | **Tuberculosis** | **Controls** | **FinnGen R7** |
| --- | --- | --- | --- |
| **N-number** | 1,895 | 307,259 | 309,154 |
| **Age of onset (mean)** | 46.56 (SD = 18.71) |  |  |
| **Sex (male)** | 1,118 (59.0%) | 134,290 (43.7%) | 135,408 (43.8%) |
| **BMI (mean)** | 26.47 (SD = 4.93) | 27.36 (SD = 5.42) | 27.35 (SD = 5.41) |

**Figure 1.**
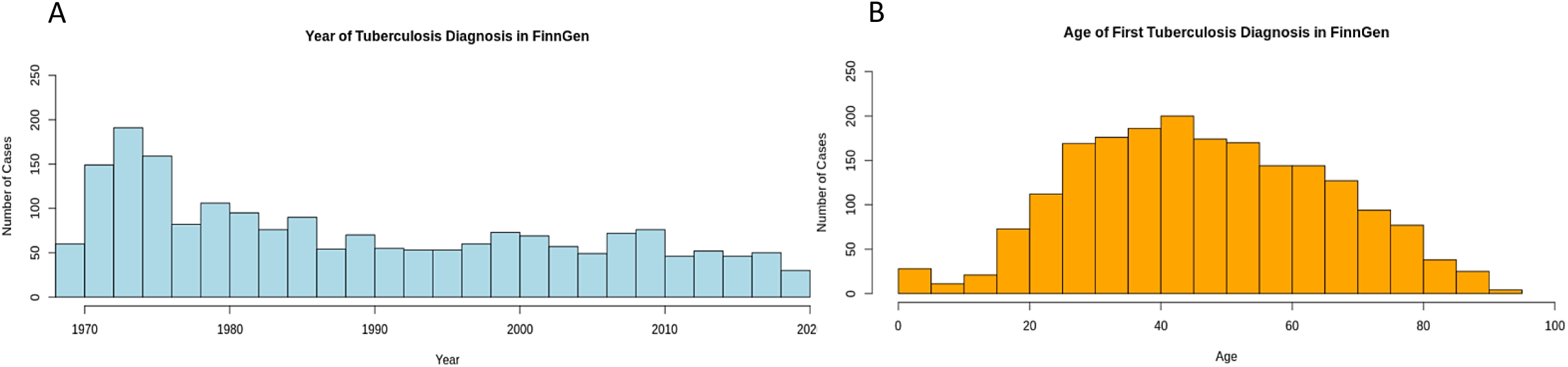
FinnGen R7 Distribution of Tuberculosis Diagnosis and Age at First Diagnosis. A: A bar chart illustrating diagnosed tuberculosis cases by year of diagnosis in FinnGen R7 starting from 1970 (start of the Care Register for Health Care Inpatient Visits in Finland) until the end of 2019 (end of follow up time 12/31/2019). B: A bar chart illustrating diagnosis age in individuals with tuberculosis (age range is from birth to 100 years-of-age).

**Figure 2.**
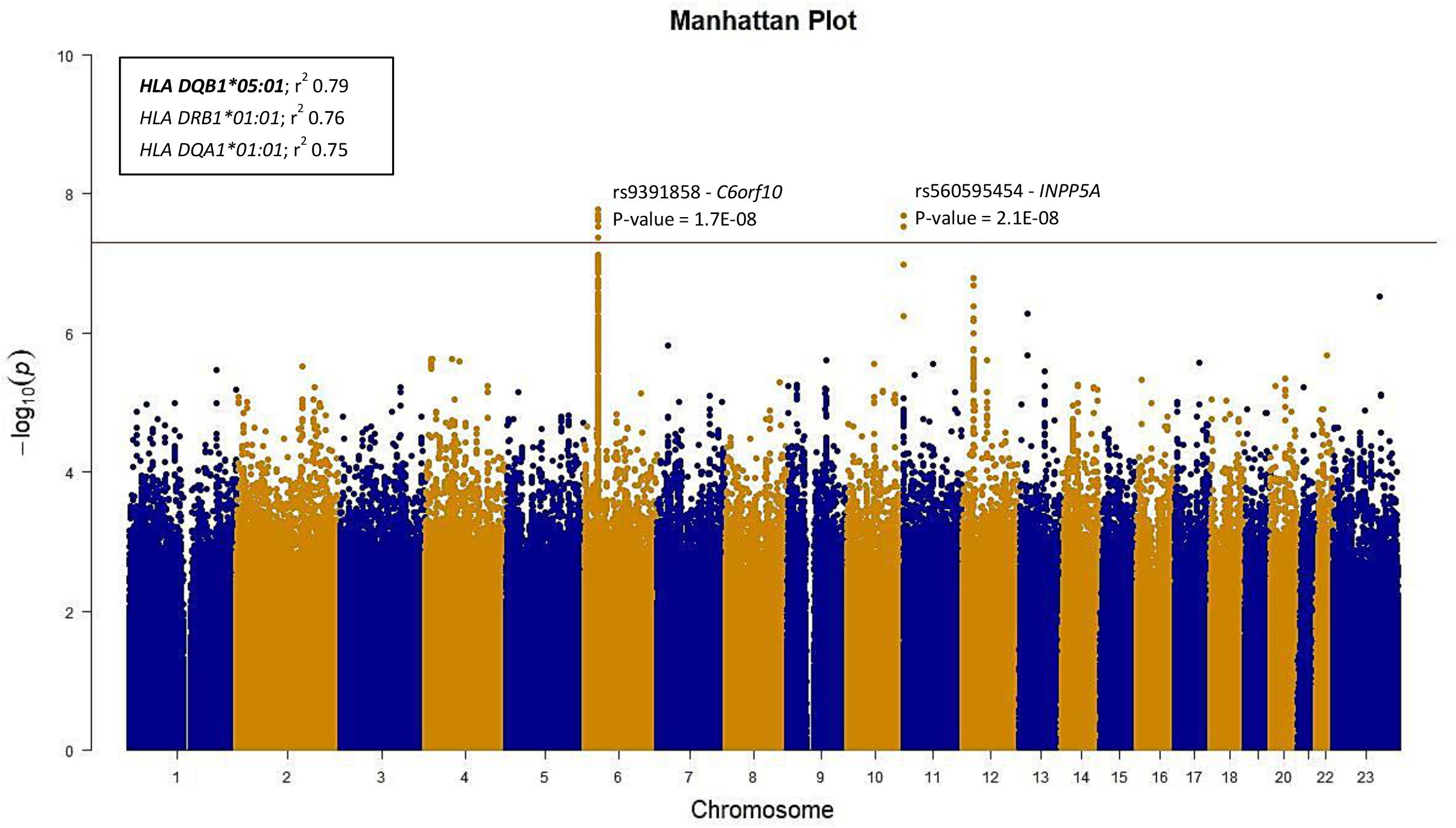
A Manhattan Plot from a Tuberculosis GWAS in FinnGen R7. A Manhattan plot from GWAS in 1,895 individuals with ICD-based tuberculosis and 307,259 controls from FinnGen R7 where each dot represent a SNP from chromosome 1 to 23. The lead variants, rs9391858 and rs560595454, and their respective P-values are marked to their corresponding positions. r^2^ values represent the linkage disequilibrium (LD) between the lead variant (rs9391858) and the respective HLA alleles.

In previous studies other genes, such as *ASAP1, IL9, WT1, ADAM12* and *MFN2*, have been shown to associate with TB (Thye *et al*., 2012; Curtis *et al*., 2015; Sobota *et al*., 2017; Qi *et al*., 2017; Quistrebert *et al*., 2021). In the FinnGen R7 cohort, we were able to replicate all the earlier reported risk variants with P<0.01 level (rs77668120 (p=6.3E-05), rs1403525097 (p=4.4E-03), rs180917274 (p=6.7E-03), rs145176301 (p=7.7E-04) and rs7530770 (p=8.1E-03), respectively). Similarly, we replicated all of the previously reported variants that have been reported in European ancestry TB GWAS (rs557011 (p=3.1E-05), rs9271378 (p=8.5E-06) and rs2894257 (p=2.6E-02)).

In this table we report the number of tuberculosis (all diagnosis subcodes) in FinnGen cohort R7, number of controls and the total number of individuals in FinnGen R7. In addition, mean age of first onset of tuberculosis, number of males in cases, controls and in FinnGen R7, and mean body mass index (BMI) are reported.

### HLA Fine Mapping

Traditionally, alleles at HLA Class II are related to transplantation outcomes, autoimmune and infectious diseases. Therefore, we used imputed HLA allele information to assess if alleles in addition to individual variants associate with TB using multivariate logistic regression adjusting for age at death or at end of follow up (12/31/2019), sex and the first 20 genetic principal components. We found three associations of protective effect estimate with HLA alleles: *HLA DQB1*05:01* (p=1.82E-06, OR=0.81 [CI 95 % 0.74-0.88]), *HLA DRB1*01:01* (p=6.86E-06, OR=0.82 [CI 95 % 0.75-0.89]) and *HLA DQA1*01:01* (p=1.004E-05, OR=0.82 [CI 95 % 0.75-0.90]) (Figure 3, Table S1-S2). Due to the high allelic diversity and high linkage disequilibrium (LD) at the HLA region (Gregersen, 2012) we measured the LD between lead variant rs9391858 at the HLA locus and individual HLA-alleles using r^2^ measure (Hill and Robertson, 1968). HLA-alleles *HLA DQB1*05:01, HLA DRB1*01:01* and *HLA DQA1*01:01* were in high LD with the lead SNP rs9391858 (r^2^=0.79, 0.76 and 0.75, respectively) (Figure 2). To test if the signal was independent we computed conditional association statistics adjusting for the lead variant rs9391858. *HLA DQB1*05:01, HLA DRB1*01:01* and *HLA DQA1*01:01* were not significant after conditioning for the lead SNP suggesting that the lead SNP and the HLA-alleles reflect the same signal (Table S3). Furthermore, we repeated the multivariate logistic regression analysis with the respiratory TB phenotype and the lead alleles remained the same (Table S4).

**Figure 3.**
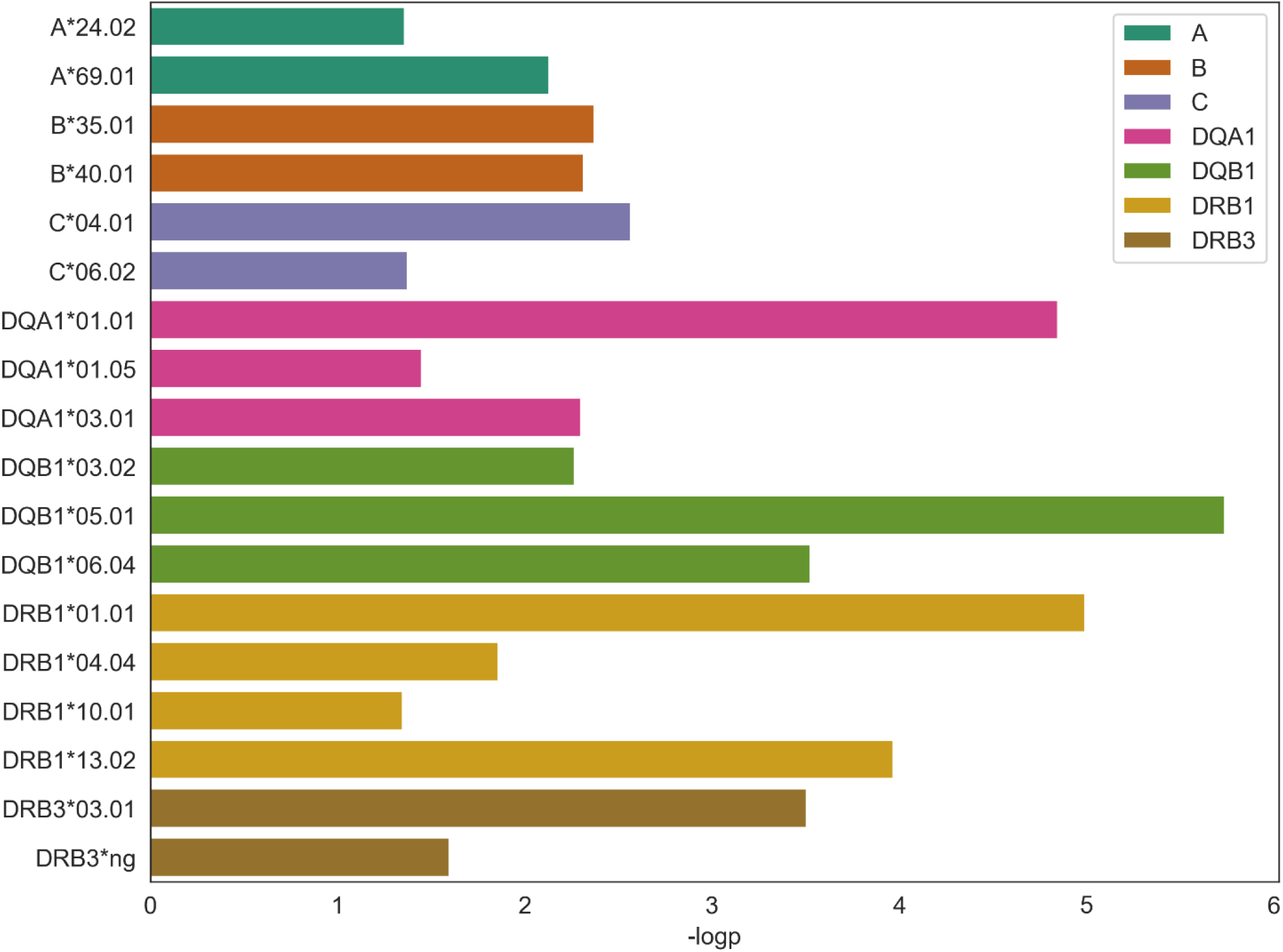
HLA Fine-mapping. Association of HLA alleles with tuberculosis in FinnGen R7 (cut off p-value 0.05).

In addition, we identified three HLA alleles that show evidence of increasing TB risk: *HLA DRB1*13:02* (p=1.07E-04, OR=1.35 [CI 95 % 1.16-1.57]), *HLA DQB1*06:04* (p=2.98E-04, OR=1.34 [CI 95 % 1.14-1.56]) and *HLA DRB3*03:01* (p=3.13E-04, OR=1.33 [CI 95 % 1.14-1.56]) (Table S2). Unlike *HLA DQB1*05:01*, the risk-alleles were not in high LD with risk SNP rs9391858 (r^2^ = 0.2) or with *HLA DQB1*05:01*. After step-wise logistic regression adjusting the HLA associations with the lead allele *HLA DQB1*05:01*, these three positive effect alleles remained statistically significant (p=0.0007, 0.002, 0.002, respectively) (Table S9).

### Epidemiological and Genetic Correlates

To study the association between known risk factors and TB, we used multivariate logistic regression and adjusted for age at death or end of follow up (12/31/2019), sex, BMI and the first 10 genetic principal components. Positive correlation with TB was witnessed in current smoking status (p=2.0E-16, OR=1.94 [CI 95 % 1.73-2.18]), ever smokers (p=2.0E-16, OR=1.87 [CI 95 % 1.66-2.12]), rheumatoid arthritis (RA) (p=1.1E-05, OR=1.65 [CI 95 % 1.32-2.07]), inflammatory bowel disease (IBD) (p=4.5E-05, OR=1.71 [CI 95 % 1.32-2.21]), Crohn’s disease (p=2.0E-02, OR=2.04 [CI 95 % 1.43-2.65]), chronic obstructive pulmonary disease (COPD) (p=2.0E-16, OR=3.71 [CI 95 % 3.26-4.22]), major coronary heart disease event (CHD) (p=3.8E-03, OR=1.17 [CI 95 % 1.05-1.30]), biological medication for rheumatoid arthritis (Bio.Med.RA) (p=1.1E-02, OR=1.91 [CI 95 % 1.42-2.40]), asthma (p=2.0E-16, OR=10.97 [CI 95 % 10.75-11.19]), alcohol use disorder (AUD) (p=1.1E-15, OR=2.11 [CI 95 % 1.76-2.53]) and alcohol dependence (p=2.0E-16, OR=2.50 [CI 95 % 2.04-3.06]) (Figure 4A and Table S5).

**Figure 4.**
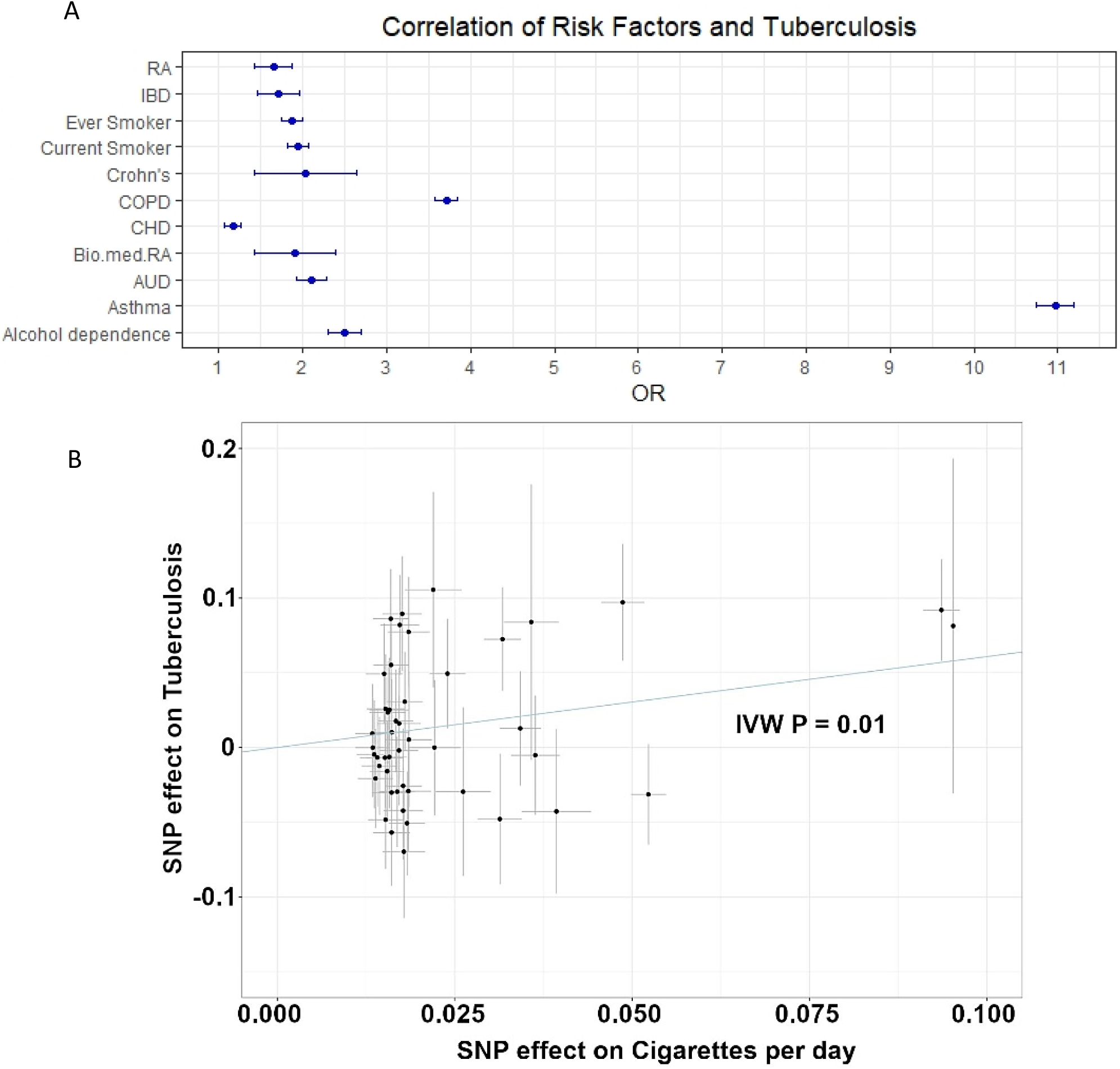
Error bar plot on association of known risk factors with tuberculosis and Mendelian randomization analysis (MR) with smoking trait. A: Bar plot demonstrates odds ratios (OR) and their respective 95 % confidence intervals between tuberculosis and risk factors. From the studied risk factors (Table S5) RA (rheumatoid arthritis), IBD (inflammatory bowel disease), ever smoker, current smoker, Crohn’s disease, COPD (chronic obstructive pulmonary disease), CHD (major coronary heart disease event), biological medication for rheumatoid arthritis (Bio.med. RA), AUD (alcohol use disorder), asthma and alcohol dependence had statistically significant and positive association with tuberculosis. B: MR suggests habitual smoking (instrumented by cigarettes per day) as a risk factor for tuberculosis (inverse-variance weighted p=0.01). The increase of habitual smoking increases the risk for tuberculosis.

To explore effect on survival we used the Kaplan-Meier estimator and Cox proportional hazards model to see the effect of TB and its known comorbidities on survival. Kaplan-Meier estimator indicated shorter lifespan within TB cases that either are current smokers or have COPD, alcohol dependence or AUD diagnosis (Figure S5-S6, Figure S8-S9). Furthermore, Cox proportional hazards model showed associations between survival and current smoking status (p=3.27E-07, HR=2.46 [CI 95 % 1.74-3.48]), COPD (p=8.98E-10, HR=1.73 [CI 95 % 1.45-2.06]), alcohol dependence (p=4.20E-07, HR=2.24 [CI 95 % 1.64-3.06]) and alcohol use disorder (p=1.45E-06, HR=2.03 [CI 95 % 1.52-2.71]) within TB patients (Table S6). Other tested comorbidities did not have an effect on survival within TB patients (Table S6). In addition, Kaplan-Meier estimator indicated shorter median survival time in non-smoking tuberculosis patients compared to smoking patients (Figure S7). We recognize that the FinnGen study may be enriched for clinical cases and effects with comorbidities may not reflect the association at population level. We therefore computed subset analysis in population sample subset of the FinnGen study (FINRISK samples: https://thl.fi/en/web/thlfi-en/research-and-development/research-and-projects/the-national-finrisk-study) that reflect better the population structure of Finland. We observed with Cox proportional hazards model that in this population smoking (ever smoker) affected the survival (p=2.00E-16, HR=1.76 [CI 95 % 1.64-1.89]) as did COPD (p=2.00E-16, HR=2.04 [CI 95 % 1.84-2.26]), alcohol dependence (p=2.00E-16, HR=3.22 [CI 95 % 2.83-3.65]) and AUD (p=2.00E-16, HR=3.05 [CI 95 % 2.74-3.41]), but TB did not (p=0.12). Furthermore, we did not see significant interaction between smoking and TB, COPD and TB, alcohol dependence and TB or AUD and TB on survival (P-value interaction 0.85, 0.42, 0.47 and 0.59, respectively).

In addition to association studies, we estimated the genetic overlap between TB and the risk factors identified from epidemiological analysis using LD score regression. In agreement with the epidemiological associations, smoking measured as number of cigarettes previously smoked daily (r_g_=0.4377, p=0.003) and current tobacco smoking (r_g_=0.3476, p=0.0048) were positively associated with TB (Table S8, Table S10).

To explore causality between TB and different traits we performed MR analysis. MR suggested smoking as a risk factor for TB (inverse-variance weighted p=0.01, OR=1.83 [CI 95 % 1.15-2.93]) with no significant pleiotropy found (Egger intercept pleiotropy test p=0.46) (Figure 4B). In addition, we tested known risk factors, BMI, vitamin D deficiency, COPD and AUD with TB, but found no causality between these traits in the FinnGen R7 cohort.

## Discussion

In this paper, we identified genetic variants from the HLA-region that protect from TB. In addition, we identified association with comorbidities, where smoking and alcohol dependence, in particular, associated with TB. Our results indicate a unique interplay between host genetic component, primarily from the *HLA DQB1*05:01* as a protective factor, and *HLA DRB1*13:02* as susceptibility factor to TB. In addition, our findings highlight the environmental component from lifestyle and comorbid factors including smoking and alcohol dependence with TB susceptibility and survival.

Our findings align with previous studies showing the association between HLA class II region and TB (See supplemental information for more detailed description) (Goldfeld *et al*., 1998; Terán-Escandón *et al*., 1999; Ravikumaret *et al*., 1999; Amirzargar *et al*., 2004; Kim *et al*., 2005; Sveinbjornsson *et al*., 2016; Oliveira-Cortez *et al*., 2016; Qi *et al*., 2017; Tian *et al*., 2017; Toyo-Oka *et al*., 2017; Bhattacharyya, Majumder, and Pandit, 2019; Tang *et al*., 2019; Seedat *et al*., 2021; Li *et al*., 2021). Other genes shown to be associated with TB susceptibility in previous studies, such as *ASAP1* (Thye *et al*., 2012), did not reach genome-wide significance in our study. In addition to HLA class II associations in TB, a recent integrative genomic analysis combining different data sets identified overall 26 candidate genes associated with TB susceptibility (Xu *et al*., 2020). This earlier evidence and our findings indicate that host and pathogen genetic factors affect disease susceptibility and severity.

There is clear heterogeneity between HLA class II allele association in different populations and different lead variants associate in different countries or ethnic groups. The reason behind the heterogeneity is unknown but may be due to selection, a bottleneck effect, pathogen driven diversity or altering virulence of different lineages of *Mycobacterium tuberculosis* bacteria (Solberg *et al*., 2008; Ejsmond and Radwan, 2011; Coscolla and Gagneux, 2014; Gagneux, 2018; Manczinger *et al*., 2019). Furthermore, HLA alleles show diversity and individual allele frequencies differ across populations, which affects the power to observe association in different populations for those alleles that are less common. In functional studies, the role of HLA class II genes in TB has started to spark interest. Kust *et al*. (2021) showed increased level of natural killer cells expressing *HLA DR* gene in the blood of primary diagnosed TB. Furthermore, Tippalagama *et al*. (2021) found that the number of *HLA DR* positive circulating CD4 T-cells was increased in patients with active form of TB. The *HLA DRB1* gene specifically has been suggested to modulate cytokine responses to *Mycobacteria tuberculosis* and thus modulate the immune response of an individual to the infection (Selvaraj *et al*., 2007).

In addition to HLA class II association with TB, we report the contribution of smoking and alcohol dependence to TB. Smoking is already a well-established risk factor in TB (reviewed in Jiang, Chen and Xie, 2020). Our results not only show a significant epidemiological association with TB, smoking habits and smoking related disease (COPD), but also show genetic correlation and causality between these traits. Through MR we identified causal relationship where increase in habitual smoking increases the risk for TB. In the FinnGen cohort TB patients were enriched for smoking throughout different decades starting from the 1970’s (Table S7). All of these gained results indicate that smoking is a major risk factor for TB, alongside with severe alcohol usage.

Our study does have some limitations. The lead SNP (rs9391858) identified by our GWAS was located nearest to the gene *HLA DRA* among the HLA genes. However, in the HLA fine mapping analysis *HLA DRA* was not among the imputed HLA genes in FinnGen. We used LD score regression to estimate the genetic correlation between different traits and TB. Our LD score regression showed low heritability within traits (1-5%), which affect the reliability of those results and therefore they should be interpreted with caution. Low heritability was most likely due to the fact that the HLA region was removed from the analysis and our results were mainly from that specific chromosomal region. Our survival analysis for epidemiological traits and TB was conducted using a Cox proportional hazards model that assumes all used traits being constant over time. We validated our Cox model evaluating the proportionality of the used predictors against time. The results showed slight statistical significance (p = 0.04), which indicates that not all predictors met the proportional assumption of the Cox model, with smoking status being one of the most evident one from the included predictors (smoking p = 0.039) (Table S6, Global p-value). Furthermore, Cox proportional hazards model only within TB patients can introduce collider bias and therefore the results should be interpreted with that kept in mind. Nevertheless, smoking was associated with TB in our causality estimates and risk factor correlations that highlights smoking being a significant risk factor in TB. Our endpoints in FinnGen for TB were defined using ICD-code based diagnoses (Tuberculosis: ICD-10 codes A15-A19; Respiratory tuberculosis: A15-A16; Tuberculosis of other organs: A17-A19). Unfortunately, we did not have information on individuals among controls who would have been infected by *Mycobacterium Tuberculosis* and might suffer from an undiagnosed latent TB infection. Additionally, we assumed in our analysis BMI and smoking information to remain unchangeable throughout the studied period due to the longitudinal nature of the data used, which does not necessarily represent the BMI and smoking information at the time of the TB diagnoses.

In Finland, cases of TB have gradually decreased from the mid-20th century to present day (Vuento, 2020). Almost half of the TB cases in Finland (year 2018) are witnessed among immigrants and transmission of TB is rare between immigrants and Finnish-born individuals (Vuento, 2020; Räisänen *et al*., 2016; Räisänen *et al*., 2021). Most of the Finnish-born cases encountered present day are elderly individuals with reactivation of a latent TB originally acquired during their childhood (Vuento, 2020). In our study, FinnGen participants were matched against a Finnish reference panel, which highlights our genetic findings to be specific to individuals of Finnish ancestry and can be also regarded as limiting factor in our study. Furthermore, it has been previously shown that individuals with rheumatoid arthritis in Finland have a higher incidence of TB compared to the general Finnish population (Vuorela *et al*., 2019). This was also witnessed in the FinnGen cohort alongside other comorbidities and risk factors that are known risk factors for TB in other populations as well (reviewed in Marais *et al*., 2013).

Our results highlight the importance of host genetic factors in TB alongside environmental risk factors. These additional results may benefit in the research and, ultimately, clinical interventions both for TB and other infectious diseases. However, further epidemiological and functional studies are needed to reveal the biological mechanisms underlying the individual reaction we have as humans to different infections.

## Data Availability

Data and code used in this study are available upon reasonable request. The FinnGen individual level data may be accessed through applications to the Finnish Biobanks FinnBB portal, Fingenious (www.finbb.fi). Summary data can be accessed through the FinnGen site https://www.finngen.fi/en/access_results.

## Supplemental data

Supplemental data include nine figures and ten tables of which two tables are in a Microsoft Office™ Excel-format. In addition, description of previous HLA-allele findings in TB and contributors of the FinnGen study are included in the supplemental data.

## Acknowledgments

We want to acknowledge the FinnGen study and the FinnGen team for their contribution. Additionally, we want to acknowledge Professor Markku Partinen (Helsinki Sleep Clinic), Dr Matti Pirinen (FIMM) and Dr Bryan Bryson (MIT) for their excellent comments and thoughts on our study design. This work was funded by the Instrumentarium Science Foundation, Signe and Ane Gyllenberg foundation and Yrjö Jahnsson foundation.

The FinnGen project is funded by two grants from Business Finland (HUS 4685/31/2016 and UH4386/31/2016) and the following industry partners: AbbVie Inc., AstraZeneca UK Ltd, BiogenMA Inc., Bristol Myers Squibb, Genentech Inc., Merck Sharp Dohme Corp, Pfizer Inc., Glaxo-SmithKline Intellectual Property Development Ltd., Sanofi US Services Inc., Maze TherapeuticsInc., Janssen Biotech Inc, Novartis Pharma AG, and Boehringer Ingelheim. Following biobanks are acknowledged for delivering biobank samples to FinnGen: Auria Biobank (www.auria.fi/biopankki), THL Biobank (www.thl.fi/biobank), Helsinki Biobank (www.helsinginbiopankki.fi), Biobank Borealis of Northern Finland (https://www.ppshp.fi/Tutkimus-ja-opetus/Biopankki/Pages/Biobank-Borealis-briefly-in-English.aspx), Finnish Clinical Biobank Tampere (www.tays.fi/en-US/Research/and/development/Finnish/Clinical/Biobank/Tampere), Biobank of Eastern Finland (www.ita-suomenbiopankki.fi/en), Central Finland Biobank (www.ksshp.fi/fi-FI/Potilaalle/Biopankki), Finnish Red Cross Blood Service Biobank (www.veripalvelu.fi/verenluovutus/biopankkitoiminta) and Terveystalo Biobank (www.terveystalo.com/fi/Yritystietoa/Terveystalo-Biopankki/Biopankki/). All Finnish Biobanks are members of BBMRI.fi infrastructure (www.bbmri.fi) and FINBB biobank cooperative (https://finbb.fi/) is the coordinator of the BBMRI-ERIC operations in Finland covering all Finnish biobanks.

## Data and code availability

Data and code used in this study are available upon reasonable request. The FinnGen individual level data may be accessed through applications to the Finnish Biobanks’ FinnBB portal, Fingenious (www.finbb.fi). Summary data can be accessed through the FinnGen site https://www.finngen.fi/en/access_results.

