## Supplemental Information for "Large Registry Based Analysis of Genetic Predisposition to Tuberculosis Identifies Genetic Risk Factors at HLA"

### Supplements

#### Table of Contents

Supplemental Figures

- Figure S1 – Locus zoom plot, tuberculosis all organs (*C6orf10*)
- Figure S2 – Locus zoom plot, tuberculosis all organs (*INPP5A*)
- Figure S3 – Manhattan plot, respiratory tuberculosis
- Figure S4 – Manhattan plot, tuberculosis of other organs
- Figure S5 – Kaplan-Meier estimate, smoking
- Figure S6 – Kaplan-Meier estimate, COPD
- Figure S7 – Kaplan-Meier estimate, smoking among tuberculosis cases and controls
- Figure S8 – Kaplan-Meier estimate, alcohol dependence
- Figure S9 – Kaplan-Meier estimate, AUD

Supplemental Tables

- Table S1 – Lead HLA-allele frequencies in FinnGen R7
- Table S2 – HLA fine mapping, tuberculosis all organs
- Table S3 – rs9391858 adjusted HLA fine mapping, tuberculosis all organs
- Table S4 – HLA fine mapping, respiratory tuberculosis
- Table S5 – Epidemiological correlates
- Table S6 – Cox proportional hazards model
- Table S7 – Changes of smoking over the decades
- Table S8 – Genetic Correlation TB vs. AUD

Supplemental Information

- Previous HLA findings from the literature
- Contributors of FinnGen

Large Registry Based Analysis of Genetic Predisposition to Tuberculosis
Identifies Genetic Risk Factors at HLA
Supplements

### Supplemental Figures

#### Figure S1 – Locus zoom plot, tuberculosis all organs (*C6orf10*)

Locus zoom plot for FinnGen R7, Tuberculosis all organs (chromosome 6, from 32.20 to 32.60).

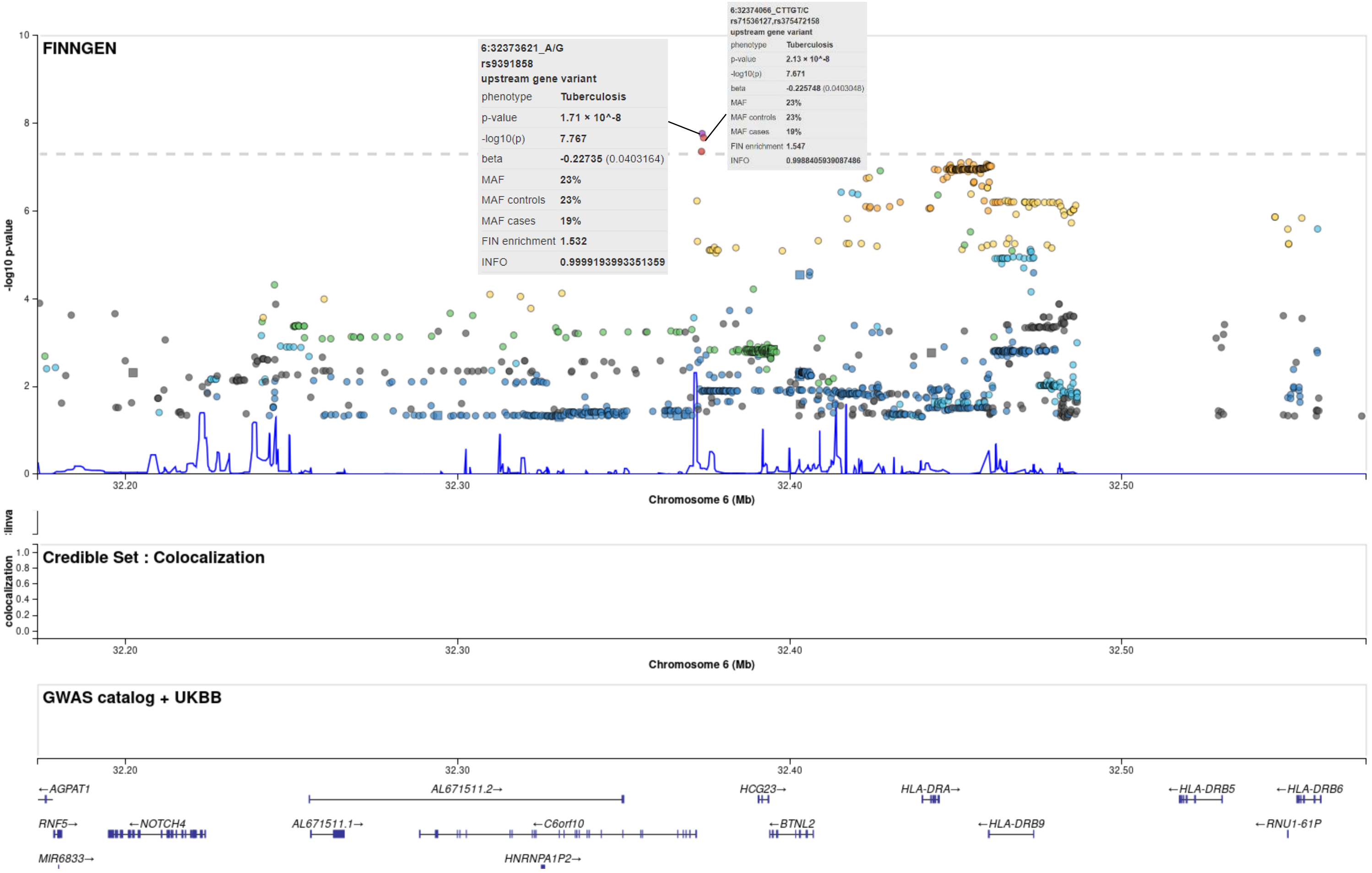

### Figure S2 – Locus zoom plot, tuberculosis all organs (*INPP5A*)

Locus zoom plot for FinnGen R7, Tuberculosis all organs (chromosome 10, from 132.50 to 132.90).

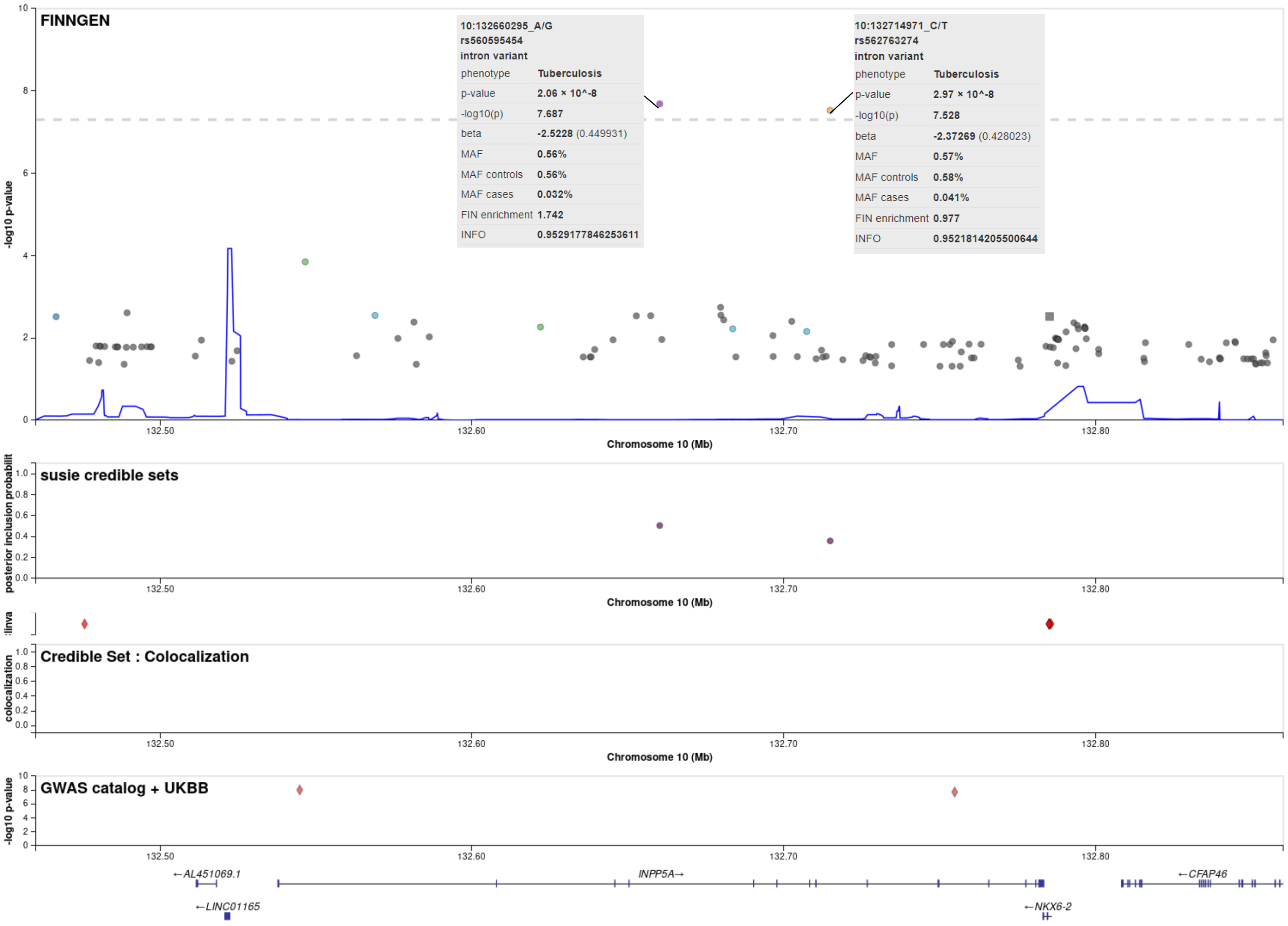

Large Registry Based Analysis of Genetic Predisposition to Tuberculosis
Identifies Genetic Risk Factors at HLA
Supplements

#### Figure S3 – Manhattan plot, respiratory tuberculosis

Manhattan Plot from FinnGen R7 GWAS – Respiratory tuberculosis (ICD-10 A15 – A16).

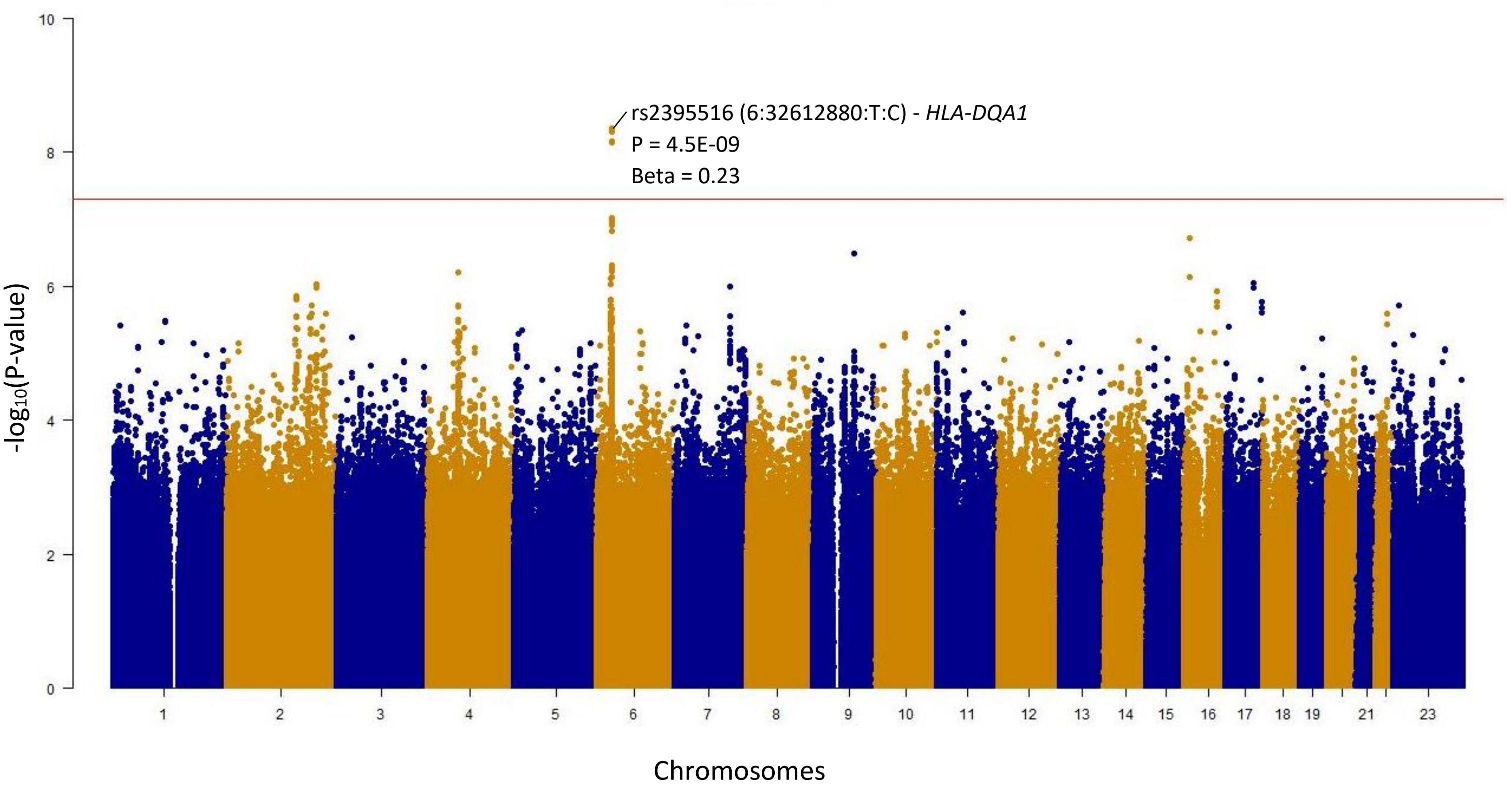

#### Figure S4 – Manhattan plot, tuberculosis of other organs

Manhattan Plot from FinnGen R7 GWAS – Tuberculosis of other organs (ICD-10 A18).

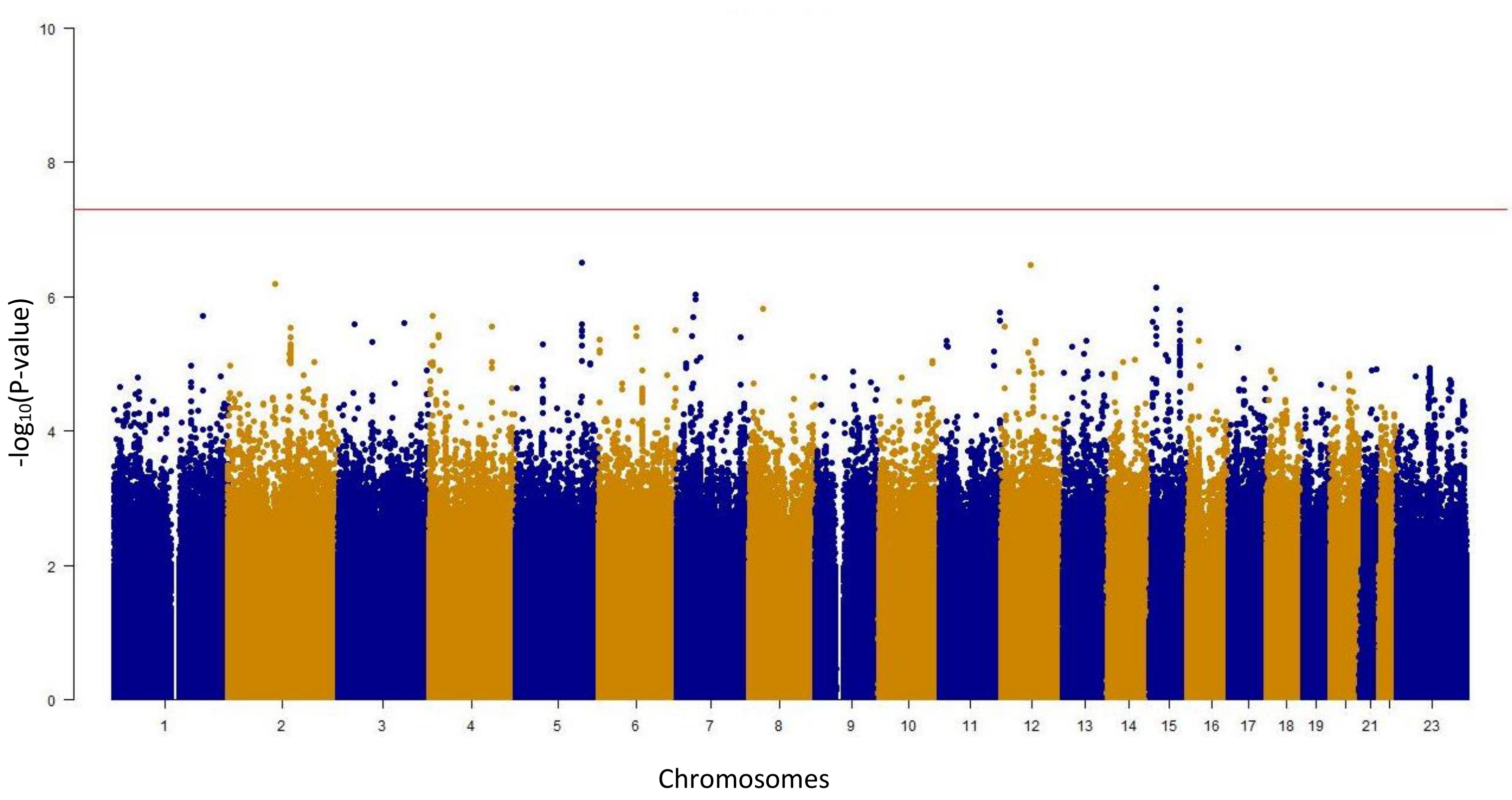

Large Registry Based Analysis of Genetic Predisposition to Tuberculosis
Identifies Genetic Risk Factors at HLA
Supplements

### Figure S5 – Kaplan-Meier estimate, smoking

A Kaplan-Meier estimate for smoking (current, former, never) among tuberculosis cases in FinnGen R7.

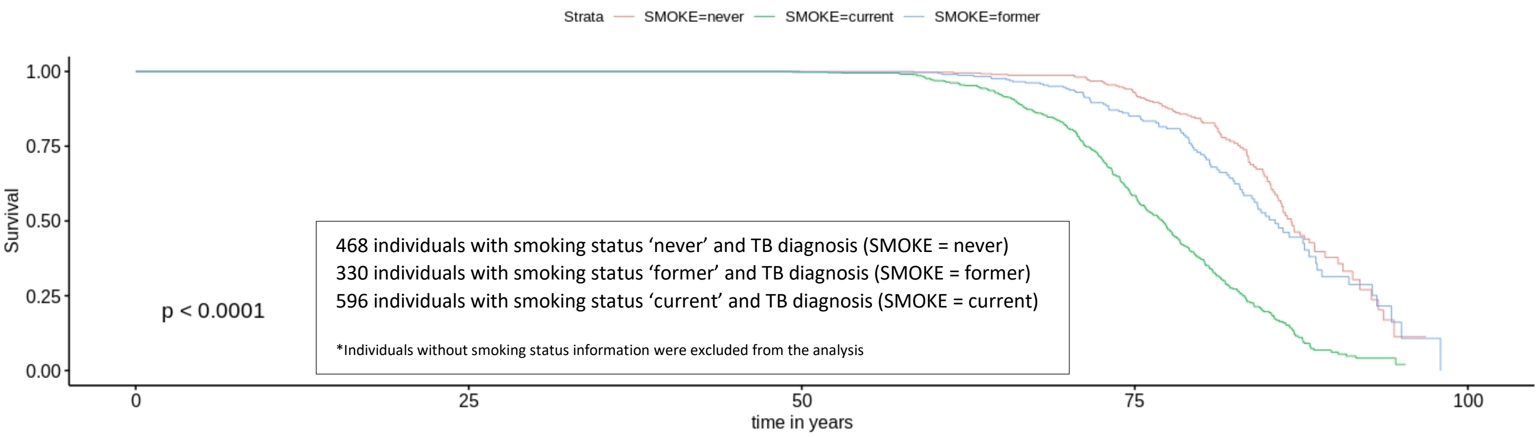


### Figure S6 – Kaplan-Meier estimate, COPD

A Kaplan-Meier estimate for COPD among tuberculosis cases in FinnGen R7.

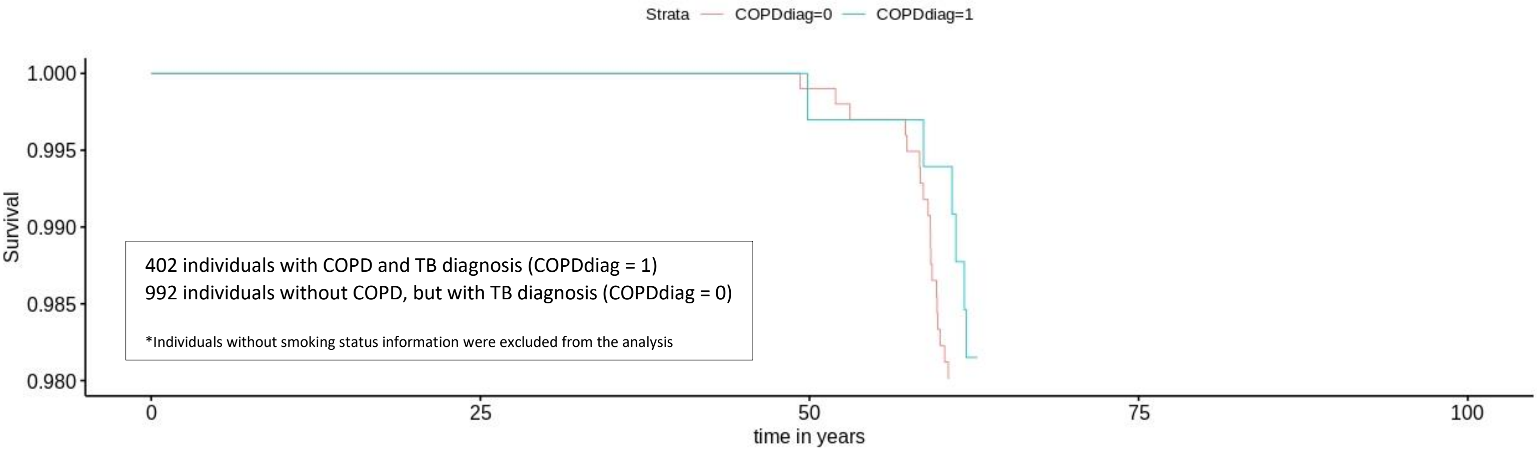


### Figure S7 – Kaplan-Meier estimate, smoking among tuberculosis cases and controls

A Kaplan-Meier estimate for smoking among tuberculosis cases and controls in FinnGen R7.

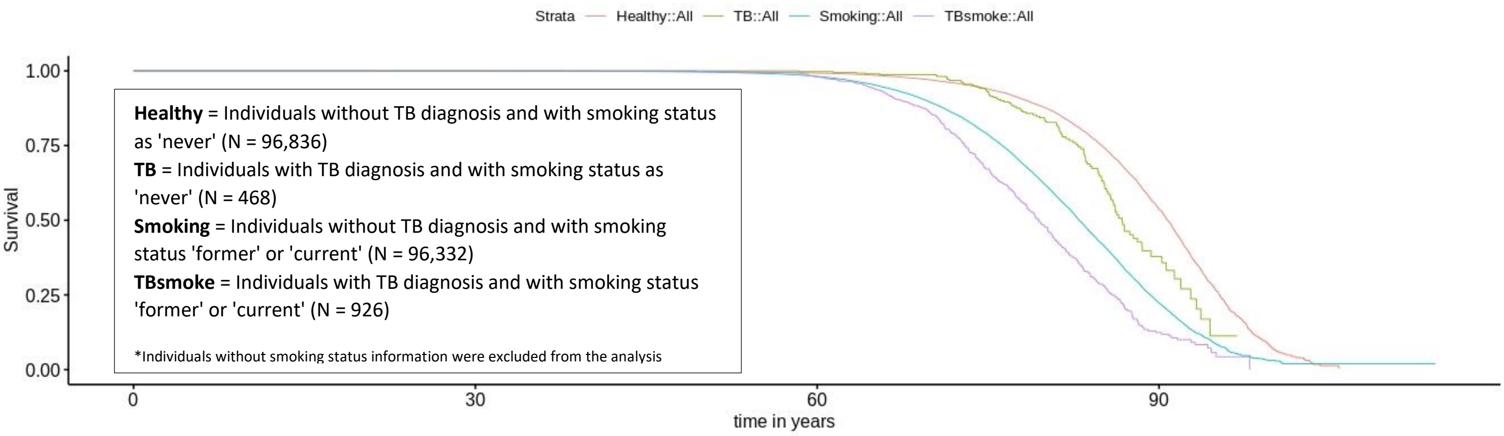

#### Figure S8 – Kaplan-Meier estimate, alcohol dependence

A Kaplan-Meier estimate for alcohol dependence among tuberculosis cases in FinnGen R7.

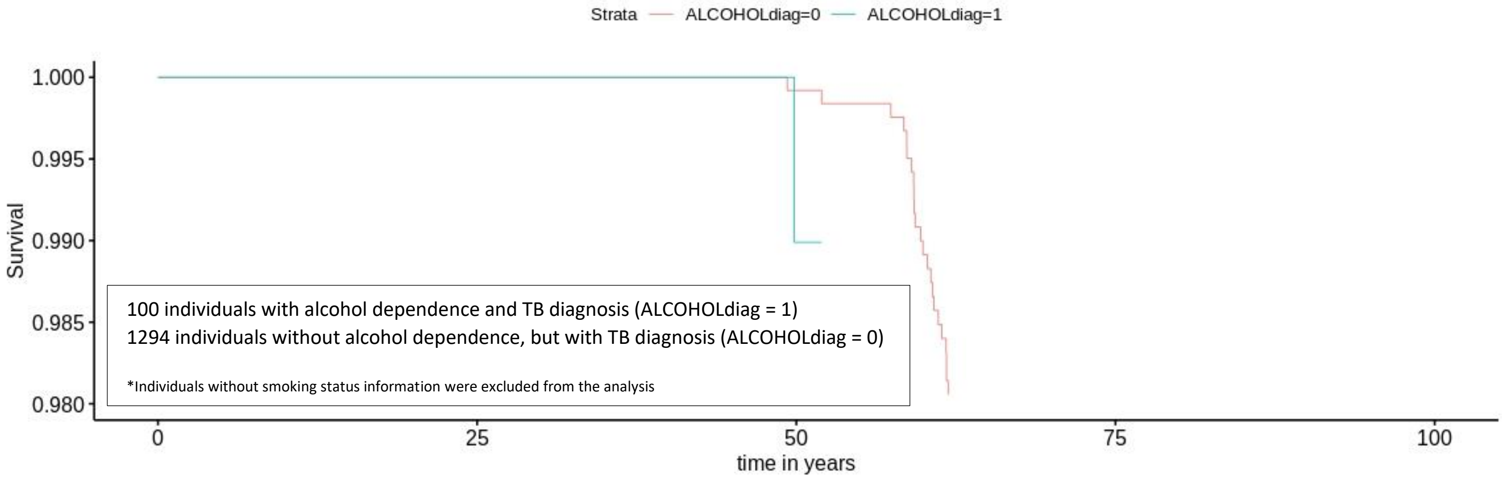


#### Figure S9 – Kaplan-Meier estimate, AUD

A Kaplan-Meier estimate for alcohol use disorder (AUD) among tuberculosis cases in FinnGen R7.

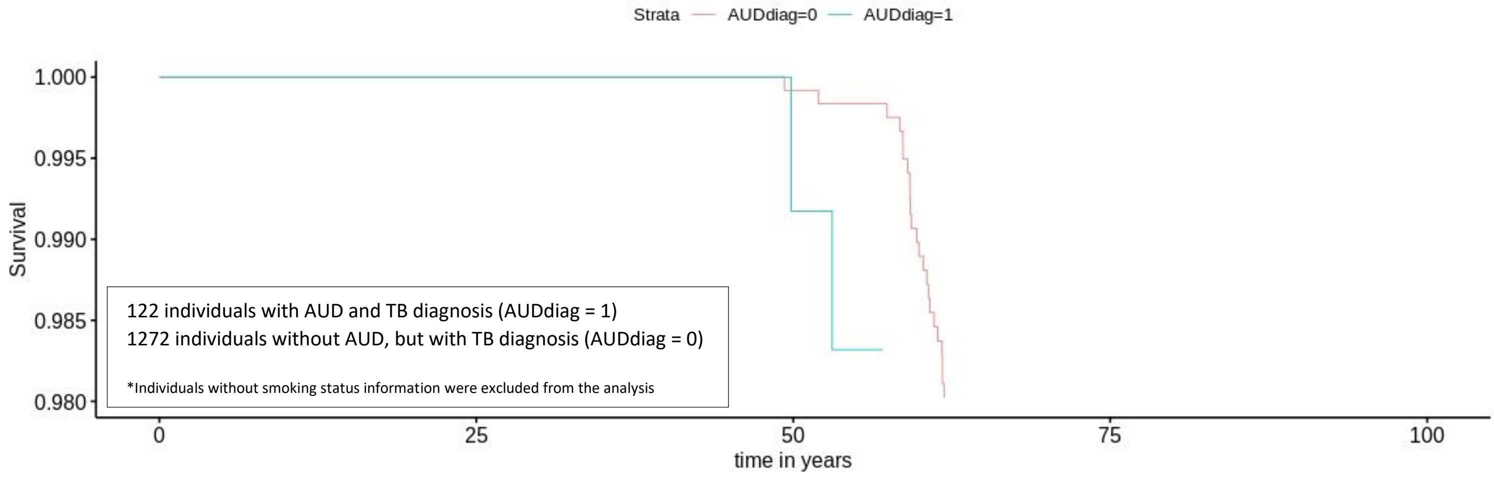

Large Registry Based Analysis of Genetic Predisposition to Tuberculosis
Identifies Genetic Risk Factors at HLA
Supplements

### Supplemental Tables

#### Table S1 – Lead HLA-allele frequencies in FinnGen R7

| HLA allele | TB AF % | CTRL AF % |
|---|---|---|
| *HLA A*24:02* | 10.08 % | 9.51 % |
| *HLA B*35:01* | 11.61 % | 13.06 % |
| *HLA B*40:01* | 7.23 % | 6.16 % |
| *HLA C*04:01* | 14.17 % | 7.68 % |
| *HLA C*06:02* | 5.33 % | 6.31 % |
| *HLA DQA1*01:01* | 15.94 % | 18.55 % |
| *HLA DQA1*03:01* | 13.06 % | 11.97 % |
| *HLA DQB1*03:02* | 13.11 % | 12.04 % |
| *HLA DQB1*05:01* | 16.49 % | 19.41 % |
| *HLA DRB1*01:01* | 15.65 % | 18.29 % |
| *HLA DRB1*13:02* | 4.70 % | 3.49 % |
| *HLA DRB3*03:01* | 4.51 % | 3.40 % |
| *HLA DRB3*ng* | 66.36 % | 68.19 % |

TB = Tuberculosis cases (ICD-10 A15-A19, all organs)
CTRL = Controls
AF = Allele frequency

NB! HLA alleles *HLA A*69:01, HLA DQA1*01:05, HLA DQB1*06:04, HLA DRB1*04:04* and *HLA DRB1*10:01* are not included in the list due to small number of carriers (< 5) of these alleles either in cases or in controls.

### Table S2 – HLA fine mapping, tuberculosis all organs

HLA fine mapping results for tuberculosis all organs (ICD-10 A15-A19).

| HLA allele | P-value | Beta (Effect Estimate) | Standard Error |
|---|---|---|---|
| *DQB1*05.01* | 1.8177189846562e-06 | -0.210785716759442 | 0.0441648730625746 |
| *DRB1*01.01* | 1.01702939035113e-05 | -0.199290409784042 | 0.0451545191592972 |
| *DQA1*01.01* | 1.41710006606709E-05 | -0.194554022988166 | 0.0448157779869284 |
| *DRB1*13.02* | 0.000107286304524348 | 0.299219345601504 | 0.0772479292295304 |
| *DQB1*06.04* | 0.00029820455831604 | 0.289068329462068 | 0.0799225712698154 |
| *DRB3*03.01* | 0.000313335622836912 | 0.283935145425041 | 0.0787829482983182 |
| *C*04.01* | 0.00270531772584413 | -0.140996097980396 | 0.0470084515624979 |
| *B*35.01* | 0.00425177392300513 | -0.146526869103373 | 0.051253748095612 |
| *B*40.01* | 0.00484644078953296 | 0.178064366825706 | 0.0632091335010802 |
| *DQA1*03.01* | 0.00500650402138516 | 0.136282593653228 | 0.0485576376432905 |
| *DQB1*03.02* | 0.00541320738891153 | 0.13480724092628 | 0.0484681441813859 |
| *A*69.01* | 0.00739186020501689 | 1.03124537853396 | 0.384986322051669 |
| *DRB1*04.04* | 0.0138169995916748 | 0.18213743461357 | 0.0739798773462318 |
| *DRB3*ng* | 0.0253384452603355 | -0.0774417927088991 | 0.0346309239484675 |
| *DQA1*01.05* | 0.0355334653287734 | -0.46213894767361 | 0.219833120984043 |
| *C*06.02* | 0.0423532666663231 | -0.147840892698834 | 0.0728268708927966 |
| *A*24.02* | 0.0438888270377818 | 0.109555253234161 | 0.054365777927801 |
| *DRB1*10.01* | 0.0449243564622073 | -0.430662534906444 | 0.214755503338578 |
| *C*03.04* | 0.0503943244175115 | 0.0931543618545088 | 0.0476102855605164 |
| *B*35.03* | 0.0763151970525559 | 0.735130245967721 | 0.414747110187033 |
| *DRB4*01.03* | 0.0782819133726585 | 0.0687675319073265 | 0.0390559737551754 |
| *DRB1*13.01* | 0.0844820220549551 | 0.0916203895218929 | 0.0531055033859619 |
| *DQA1*04.01* | 0.0879607650802239 | 0.0915010367952343 | 0.0536268498916045 |
| *DQB1*04.02* | 0.0957405982264579 | 0.0893435524736166 | 0.0536319756438577 |
| *B*55.01* | 0.0975154523671616 | -0.480766751083659 | 0.290139395320536 |
| *DRB1*08.01* | 0.102865984882623 | 0.0880481369122617 | 0.053980325526577 |
| *DRB1*08.03* | 0.105056263690922 | 0.621239772186696 | 0.383287348615493 |
| *A*33.03* | 0.105984365130154 | 0.454765430035294 | 0.281325685764493 |
| *C*08* | 0.106903291585175 | 0.945571626929328 | 0.586484818168548 |
| *DPB1*14.01* | 0.118091271049969 | 0.201377353574537 | 0.128853831495685 |
| *A*26.01* | 0.122713349405695 | -0.218468144125561 | 0.141542200885544 |
| *DRB4*01.03N* | 0.122786575163452 | -0.237860213189393 | 0.154136182608343 |
| *A*68.01* | 0.1230486059813 | -0.110474000005121 | 0.0716385428035059 |
| *B*37.01* | 0.130383835621225 | -0.332431071529675 | 0.219776060478484 |
| *DRB4*ng* | 0.135393398151014 | -0.0565916418512358 | 0.0379004024979995 |
| *C*08.02* | 0.13618584316204 | -0.609093375766225 | 0.408747440230807 |
| *DQB1*06.03* | 0.136604369006502 | 0.0824553498180087 | 0.0553928498427625 |
| *B*35.08* | 0.15001645301873 | 0.4033646817409 | 0.280216844186362 |
| *DQA1*06.01* | 0.16046603648442 | 0.580503983827274 | 0.413609937434405 |
| *B*13.02* | 0.170272089636029 | -0.139878063785119 | 0.102001748552622 |
| *DPB1*02.02* | 0.203787003056003 | -1.27243301346909 | 1.00125667357903 |

| | | | |
|---|---|---|---|
| *DQA1*01.03* | 0.210908934562964 | 0.069529049518727 | 0.0555756662256134 |
| *B*39.24* | 0.212569581895187 | -1.24953623560994 | 1.00241102446185 |
| *DRB1*04.01* | 0.221705646480324 | 0.0678769574697098 | 0.0555455536546156 |
| *A*02.05* | 0.223624958533935 | 0.551136324603185 | 0.452884974626464 |
| *DQB1*06.02* | 0.236504565702422 | -0.0572322565210728 | 0.0483474922060155 |
| *DRB1*04.08* | 0.237774318773817 | 0.167979959581486 | 0.142287387857529 |
| *DRB5*01.01* | 0.264332697321114 | -0.0537207817959755 | 0.048127889500659 |
| *DRB1*15.01* | 0.275515105439859 | -0.0523998770957568 | 0.048053457019257 |
| *DRB4*01.01* | 0.27818516941114 | 0.184840234297255 | 0.170453102356511 |
| *DRB3*01.01* | 0.298122554239283 | 0.0437467503143608 | 0.0420452652945288 |
| *B*14.02* | 0.303239816108963 | -0.389976503816941 | 0.378797960652918 |
| *C*01.02* | 0.304236873238486 | 0.0639879617767474 | 0.0622820581711716 |
| *B*44.03* | 0.316182701544759 | 0.166621485265416 | 0.166233631925474 |
| *B*35* | 0.322977446270467 | -0.989503808531814 | 1.00115950589402 |
| *DQB1*06.09* | 0.330163582014789 | 0.310817977406046 | 0.319185607804994 |
| *DQA1*01.02* | 0.331786477572933 | 0.0404872458963455 | 0.0417169887736537 |
| *DPB1*11.01* | 0.336032150941425 | 0.323286422940938 | 0.336044435563007 |
| *DRB5*ng* | 0.33705943577135 | 0.0450187496394667 | 0.0468949405447581 |
| *DQA1*02.01* | 0.33991790071603 | -0.0700678023657002 | 0.0734211301879312 |
| *A*01.01* | 0.340042211367654 | -0.0560977882328971 | 0.058797668239361 |
| *A*29.02* | 0.34081578650646 | -0.338245513588967 | 0.355093043093961 |
| *DRB1*12.01* | 0.348529625433797 | -0.100361983002625 | 0.107059045524032 |
| *DRB1*07.01* | 0.356370421369975 | -0.0677396007803394 | 0.0734461263974636 |
| *B*27.05* | 0.358227760563913 | 0.054585770066764 | 0.0594132573515397 |
| *DPB1*16.01* | 0.36583093177228 | 0.189601884551215 | 0.209664649826165 |
| *DPB1*10.01* | 0.366243077116351 | -0.453765239534136 | 0.502212132482985 |
| *C*02.02* | 0.366310920555611 | 0.0613396020020389 | 0.0678982222370706 |
| *A*23.01* | 0.368787586710068 | 0.220788212463588 | 0.245662329697599 |
| *A*02.01* | 0.372185417830053 | 0.0307670629432157 | 0.0344772562765555 |
| *B*40.02* | 0.373152870459182 | -0.0944979666818142 | 0.106107972378237 |
| *DQA1*01.04* | 0.379516087124905 | 0.125781867785875 | 0.143131027098078 |
| *A*03* | 0.391736848674247 | 0.384566605772333 | 0.449012556851048 |
| *DQB1*03.03* | 0.398761513334163 | -0.0614351891480362 | 0.0728047260182709 |
| *DRB1*09.01* | 0.403918613086283 | -0.0765790775273788 | 0.0917506721290294 |
| *B*44.27* | 0.408207142218893 | 0.15040692691705 | 0.181858920174207 |
| *B*44.02* | 0.417924110260539 | 0.053185667119907 | 0.0656590515435432 |
| *C*05.01* | 0.426262924392794 | 0.0550918937187666 | 0.0692454727715279 |
| *DRB1*01.02* | 0.426680732538495 | 0.252624184502834 | 0.317812470869564 |
| *DPB1*105.01* | 0.433808991319066 | -0.351356687139019 | 0.448908949934285 |
| *B*13.01* | 0.434882607447596 | 0.00281455235196667 | 0.003604408817878 |
| *B*46.01* | 0.434882607447596 | 0.00281455235196667 | 0.003604408817878 |
| *C*04.06* | 0.434882607447596 | 0.00281455235196667 | 0.003604408817878 |
| *DRB1*07.03* | 0.434882607447596 | 0.00281455235196667 | 0.003604408817878 |
| *DRB5*01.02* | 0.434882607447596 | 0.00281455235196667 | 0.003604408817878 |
| *DQB1*05.02* | 0.445880357638814 | 0.119351167584531 | 0.156566976102728 |

| | | | |
|---|---|---|---|
| *DRB1*04.03* | 0.452787637915095 | -0.753290209483476 | 1.00334917504318 |
| *B*39.06* | 0.455167249060998 | 0.377627392059878 | 0.505641556860554 |
| *B*07* | 0.459281321482859 | 0.371276073340665 | 0.5017035592828 |
| *C*15.02* | 0.463271452298095 | -0.077080123957799 | 0.105089588927436 |
| *A*11.01* | 0.466172545127612 | -0.0611590141062414 | 0.0839265530192635 |
| *DRB1*07* | 0.469309188838795 | 0.732284089135758 | 1.01199632652673 |
| *DQB1*05.03* | 0.480477383011462 | 0.105281101116231 | 0.149221664316657 |
| *C*12.02* | 0.4939630243967 | -0.680385942500892 | 0.994688328988244 |
| *DQA1*03* | 0.500646846203985 | 0.15587062315807 | 0.23144324738839 |
| *B*27.02* | 0.509633962832876 | 0.271874141747457 | 0.412300491173334 |
| *C*07.01* | 0.520862245022935 | 0.0312972630897993 | 0.0487483518221652 |
| *DRB1*14.01* | 0.522910691652195 | 0.193669141054982 | 0.303145593896874 |
| *DRB1*04.05* | 0.523474069579875 | 0.456271614900426 | 0.715159941703594 |
| *DRB1*11.04* | 0.524702661609891 | 0.165464587933868 | 0.260118160644715 |
| *C*14.02* | 0.527574499708864 | -0.0902340371659043 | 0.142840264521325 |
| *B*35.02* | 0.565691302418146 | -0.405450632198974 | 0.705857465488323 |
| *DRB1*08.02* | 0.566129982745985 | 0.235787383163422 | 0.410950988272982 |
| *B*57.01* | 0.573939859164288 | -0.0857380904232931 | 0.152488720541377 |
| *DQB1*02* | 0.576471254415867 | -0.561215371641009 | 1.00477822682713 |
| *C*07.02* | 0.580956746628585 | 0.0251533314600913 | 0.0455686407726501 |
| *A*30* | 0.581168893267329 | -0.247447052758817 | 0.448535182495804 |
| *DRB1*11.03* | 0.582951706927963 | -0.388849795659698 | 0.70818640224644 |
| *DQA1*03.03* | 0.590646253505403 | 0.0536768818482699 | 0.0997897484796609 |
| *B*18.01* | 0.591409593108362 | 0.0395376578222373 | 0.0736551141194091 |
| *DRB5*02.02* | 0.594485050388055 | 0.0951538606746261 | 0.178743745518246 |
| *B*52.01* | 0.600121826139544 | -0.521596035068475 | 0.994984421316277 |
| *C*12.03* | 0.603453931959729 | 0.0468458369352815 | 0.0901852676720005 |
| *C*07.04* | 0.628728569501428 | 0.0560089167452999 | 0.115836458166223 |
| *DQA1*05.01* | 0.638531424200692 | -0.0258470194500426 | 0.0550225625703097 |
| *B*15.17* | 0.639220912079732 | -0.333313180415022 | 0.711009914309195 |
| *B*41.01* | 0.647245175130724 | -0.16293687922847 | 0.35607412851009 |
| *A*29.01* | 0.648676767150291 | -0.152733046619572 | 0.33523406127135 |
| *DQB1*02.01* | 0.652257352174899 | -0.0248554874965423 | 0.0551573852585544 |
| *B*58.01* | 0.663105315750908 | 0.117583745746371 | 0.269916645892529 |
| *DRB1*03.01* | 0.666014985369176 | -0.0237752110140309 | 0.055083201362533 |
| *C*17.01* | 0.673919986679523 | -0.149834626255976 | 0.356092728450365 |
| *DQA1*05.05* | 0.694599031837556 | -0.0249514297232692 | 0.0635508435366065 |
| *B*15.01* | 0.701440247699453 | 0.0184216115085211 | 0.0480509031002791 |
| *DRB1*13.03* | 0.705853994613457 | 0.087349232567967 | 0.231431542985132 |
| *DPB1*15.01* | 0.712830305724589 | 0.0850220860241691 | 0.231002137499138 |
| *C*17.03* | 0.721756956377953 | 0.119538463350768 | 0.335676928527952 |
| *DRB1*14.54* | 0.72212325503525 | 0.0598400654582647 | 0.16832476606753 |
| *DRB1*16.01* | 0.757676938687422 | 0.0542891308104996 | 0.175959020019271 |
| *B*41.02* | 0.757802055626712 | 0.103511547426305 | 0.335674995410517 |
| *DPB1*19.01* | 0.758384243473144 | -0.0750086741898393 | 0.24384876652162 |

| | | | |
|---|---|---|---|
| *C*07* | 0.780485004191064 | 0.276964642293591 | 0.99381957548765 |
| *B*14.01* | 0.7855995450811 | -0.1935038741947 | 0.711334862701405 |
| *C*03.02* | 0.792336611279078 | 0.0710552585691577 | 0.269887200088487 |
| *B*18* | 0.797807241407365 | -0.182116151969012 | 0.710874806467924 |
| *DPB1*13.01* | 0.804517429888373 | -0.0751875725570702 | 0.303782046792264 |
| *B*47.01* | 0.806017704657732 | -0.0485763263201402 | 0.197813202261949 |
| *A*30.01* | 0.815632011131466 | -0.0783576968189674 | 0.336058784310323 |
| *DQA1*03.02* | 0.818974132357188 | -0.0187089774492801 | 0.0817469150792993 |
| *A*03.01* | 0.820103613716189 | -0.00862290944025734 | 0.0379176136134433 |
| *A*31.01* | 0.827966560246977 | 0.0177833233033665 | 0.0818337944015984 |
| *A*32.01* | 0.830886067827977 | -0.0184441337579149 | 0.0863629739616395 |
| *DRB1*14.02* | 0.836961330331688 | -0.0555346942899879 | 0.269871701867855 |
| *B*39.01* | 0.83865430170205 | 0.0175707497937577 | 0.0862939479325443 |
| *DRB1*14* | 0.839716458659061 | 0.203703373189796 | 1.00715526998238 |
| *DQA1*05.03* | 0.85300629722446 | -0.0500054512736696 | 0.269885445185694 |
| *DRB1*11.01* | 0.854294787886244 | 0.0158522551427697 | 0.0863217972152752 |
| *A*25.01* | 0.86613562671491 | 0.0252169260790672 | 0.149594045235127 |
| *DPB1*17.01* | 0.88055700319142 | -0.0391684541364081 | 0.260665729262324 |
| *B*38.01* | 0.88641163282547 | 0.0268875850223321 | 0.188227486614236 |
| *B*44* | 0.891920780516935 | -0.0969333353914567 | 0.713405336933239 |
| *B*51.01* | 0.893340864521975 | 0.00914083873046681 | 0.0681755098968072 |
| *B*08.01* | 0.894088565871156 | 0.00757878418149007 | 0.0569266126229567 |
| *C*03.03* | 0.894630090473503 | -0.00778844232077018 | 0.0588038530117866 |
| *DPB1*06.01* | 0.89667429361045 | -10.4203506553276 | 80.240695705409 |
| *B*49.01* | 0.896760793235044 | 0.0585940142190326 | 0.451576546403858 |
| *B*39* | 0.898250069614947 | -10.3149437843561 | 80.6660048520113 |
| *A*33.01* | 0.903346399547031 | -11.351631455454 | 93.4789811042933 |
| *B*07.02* | 0.90338201939841 | 0.00577067503327127 | 0.0475382483369377 |
| *DRB1*01.03* | 0.904151395116224 | -10.4470522307304 | 86.7559861789091 |
| *C*04* | 0.914069649975851 | -8.42896347391847 | 78.1133916502527 |
| *DPB1*05.01* | 0.914408236070329 | -0.0117350076596214 | 0.109183232695895 |
| *DRB1*15.02* | 0.914653458600706 | -10.2054029251424 | 95.225568263416 |
| *DQB1*06.01* | 0.917648709496265 | -10.2132658082243 | 98.7781795154543 |
| *A*02.02* | 0.920117129416746 | -9.30910601370167 | 92.82541292781 |
| *DPB1*20.01* | 0.920341743786007 | -0.0225849424396553 | 0.225842080350658 |
| *A*02* | 0.920844365887646 | -10.4199766361551 | 104.860206717291 |
| *B*15* | 0.923329759651805 | -10.3203121319818 | 107.234878406431 |
| *B*13* | 0.928319688057325 | -7.91285967708988 | 87.9606013415731 |
| *B*45.01* | 0.930778898174846 | 0.0437580319084246 | 0.503748129557689 |
| *B*15.16* | 0.931612189676668 | -7.11121883713974 | 82.8652812008156 |
| *B*51* | 0.932483598987287 | 0.0381143555160991 | 0.449883512515211 |
| *DQB1*03.01* | 0.932552492770001 | 0.00442437114716164 | 0.0522766187969216 |
| *C*12* | 0.933599136432028 | -8.21427725853072 | 98.5901347341893 |
| *DQB1*03.04* | 0.933720281435844 | -9.30887284694634 | 111.932475025935 |
| *DQB1*06* | 0.934338477107547 | -7.14630795533433 | 86.7400660107676 |

| | | | |
|---|---|---|---|
| *DQB1*02.02* | 0.934964843704563 | -0.00677935016122119 | 0.08308032587934 |
| *DQB1*04* | 0.935147706857262 | -7.41004812280394 | 91.0660951552204 |
| *DPB1*04.02* | 0.935297895115271 | -0.00336965526613974 | 0.0415078565062081 |
| *A*01* | 0.935813342447317 | -7.45019600853162 | 92.5110822137331 |
| *C*01* | 0.936555801352831 | -6.20933519728987 | 78.0072239959559 |
| *C*16.02* | 0.940329212445388 | -7.11229043363161 | 95.0128479431829 |
| *A*02.17* | 0.940491370511886 | -7.76761011245015 | 104.050530028168 |
| *C*03* | 0.941529286630445 | -6.31377637401294 | 86.079868185761 |
| *A*32* | 0.941775922467487 | -6.27840905414872 | 85.9609215850848 |
| *DPB1*04.01* | 0.942378890251153 | -0.00240920473189334 | 0.0333314437345499 |
| *DRB1*15* | 0.942691139925345 | -7.10071079984918 | 98.7748053633227 |
| *C*02* | 0.944063016525347 | -5.83298270991595 | 83.1333891567483 |
| *DPB1*02.01* | 0.944613412763874 | -0.00327249465492291 | 0.0471047999657795 |
| *A*31* | 0.944703369533026 | -8.55072366411657 | 123.281003905146 |
| *DPB1*09.01* | 0.946733460686489 | -7.01257911972163 | 104.963999757383 |
| *C*06* | 0.947168875959668 | -5.52957568565128 | 83.4496078287949 |
| *A*28* | 0.947191304066621 | -6.35317535308314 | 95.9197445415956 |
| *A*11* | 0.948528091964294 | -8.0776105946131 | 125.127032562227 |
| *A*30.02* | 0.949659011012729 | -7.16330520597424 | 113.460144416911 |
| *A*01.02* | 0.95145784044147 | -7.20702001051233 | 118.388227874783 |
| *A*02.06* | 0.951681149640123 | -6.62104020793914 | 109.265732626595 |
| *A*68* | 0.951716314890193 | -6.72145155016685 | 111.003685931058 |
| *B*40* | 0.952943888945952 | -6.59418259937698 | 111.746262259295 |
| *DRB1*13* | 0.95323055295164 | -6.48234867637093 | 110.52519364611 |
| *DRB3*02.02* | 0.954587740470572 | -0.00305607509661827 | 0.0536656375312709 |
| *B*27* | 0.955188655567503 | -6.11775410947589 | 108.87184950198 |
| *B*08* | 0.955428338603025 | -5.39586317368401 | 96.5419580559696 |
| *DPB1*03.01* | 0.958495076335428 | -0.00250837352387387 | 0.0481988466095366 |
| *DRB1*04.02* | 0.958665376568694 | -4.22819948521381 | 81.5806473152724 |
| *DPB1*01.01* | 0.958862790618662 | 0.00366605528118049 | 0.0710741540872702 |
| *A*68167* | 0.959823936452089 | -5.47755984315372 | 108.736703876566 |
| *C*15.05* | 0.961948909942061 | -0.0480291060662568 | 1.00672937448654 |
| *A*24* | 0.96244045390872 | 0.0332634002971891 | 0.706359641778003 |
| *C*05* | 0.964254503621785 | -5.35400318811261 | 119.468111963819 |
| *DPB1*34.01* | 0.96430796645232 | -5.34598843683063 | 119.468074564843 |
| *B*50.01* | 0.975807607944439 | -0.0152441472277788 | 0.502687196109129 |
| *C*16.01* | 0.985346207386862 | -0.00494144245409482 | 0.269041543391394 |
| *B*56.01* | 0.986736297924451 | 0.00194007909365754 | 0.116701042316236 |
| *DRB1*04.07* | 0.999170058519066 | -0.000468264037668255 | 0.450177015179505 |

#### Table S3 – rs9391858 adjusted HLA fine mapping, tuberculosis all organs

HLA fine mapping results for tuberculosis all organs (ICD-10 A15-A19) adjusted with the lead SNP rs9391858.

| HLA allele | P-value | Beta (Effect Estimate) | Standard Error |
|---|---|---|---|
| *DRB1*13.02* | 0.001157836 | 0.252427934 | 0.077692404 |
| *DQB1*06.04* | 0.002586831 | 0.242096796 | 0.080350875 |
| *DRB3*03.01* | 0.002790512 | 0.236858687 | 0.079219092 |
| *A*69.01* | 0.009833302 | 0.994161398 | 0.385089571 |
| *DPB1*04.02* | 0.013360253 | 0.11225536 | 0.045373683 |
| *B*40.01* | 0.019551177 | 0.148099385 | 0.063429893 |
| *DQB1*06.02* | 0.019625068 | -0.114713839 | 0.049160828 |
| *DRB5*01.01* | 0.022644833 | -0.111591143 | 0.048956922 |
| *DRB1*15.01* | 0.024388635 | -0.110025335 | 0.048879487 |
| *DRB5*ng* | 0.029971531 | 0.103693787 | 0.047774887 |
| *A*68.01* | 0.056297496 | -0.137008408 | 0.071780043 |
| *A*24.02* | 0.068727923 | 0.099003528 | 0.054391442 |
| *C*06.02* | 0.069316637 | -0.132382516 | 0.072883798 |
| *B*55.01* | 0.073398173 | -0.51959099 | 0.290218064 |
| *DRB1*04.04* | 0.075104538 | 0.132577357 | 0.074488968 |
| *DQA1*03.01* | 0.086100444 | 0.084862119 | 0.049443749 |
| *B*35.03* | 0.089448031 | 0.704433684 | 0.414783499 |
| *DQB1*03.02* | 0.090068397 | 0.083640655 | 0.049344441 |
| *A*33.03* | 0.094010783 | 0.470916912 | 0.281209907 |
| *C*08.02* | 0.110644104 | -0.65213503 | 0.408783983 |
| *C*03.04* | 0.111110676 | 0.076002456 | 0.047703657 |
| *B*13.02* | 0.112169604 | -0.162163911 | 0.102085131 |
| *C*08* | 0.129128689 | 0.890313968 | 0.586679747 |
| *A*26.01* | 0.12981225 | -0.214412568 | 0.141541146 |
| *DRB1*08.03* | 0.129982301 | 0.580458019 | 0.383350201 |
| *DQA1*05.01* | 0.143667907 | -0.081505929 | 0.055739409 |
| *DQB1*02.01* | 0.149393997 | -0.080553434 | 0.05587488 |
| *DPB1*14.01* | 0.150342148 | 0.185403663 | 0.128902573 |
| *DRB1*03.01* | 0.1546372 | -0.079423898 | 0.055801015 |
| *DRB1*12.01* | 0.15759658 | -0.151760374 | 0.107387664 |
| *A*01.01* | 0.185936418 | -0.077901696 | 0.058896136 |
| *DQA1*06.01* | 0.187231072 | 0.545523733 | 0.413646994 |
| *DPB1*02.02* | 0.191759213 | -1.307025628 | 1.001251022 |
| *B*35.08* | 0.194911193 | 0.363308338 | 0.280290144 |
| *B*39.24* | 0.19724799 | -1.292573877 | 1.002436742 |
| *DQA1*01.05* | 0.202146529 | -0.283839208 | 0.222538726 |
| *A*03.01* | 0.21630984 | 0.048336242 | 0.039094334 |
| *C*01.02* | 0.217115507 | 0.076940411 | 0.062338564 |
| *B*27.05* | 0.219451254 | 0.07303542 | 0.059475651 |
| *DQA1*05.05* | 0.223167511 | -0.078124352 | 0.06413368 |

| | | | |
|---|---|---|---|
| *B*37.01* | 0.242232546 | -0.257378198 | 0.220089578 |
| *DRB1*10.01* | 0.246522141 | -0.252057365 | 0.217508677 |
| *B*14.02* | 0.250059154 | -0.435775261 | 0.378867237 |
| *A*02.05* | 0.257275778 | 0.513088764 | 0.452917136 |
| *B*40.02* | 0.288985209 | -0.112566554 | 0.106159767 |
| *DRB3*02.02* | 0.290319744 | -0.057511921 | 0.054388886 |
| *B*35* | 0.304747317 | -1.027549638 | 1.001211651 |
| *A*29.02* | 0.309394299 | -0.36095267 | 0.355095792 |
| *DQA1*01.01* | 0.338882339 | 0.088992171 | 0.093051454 |
| *DQB1*03.01* | 0.344540677 | -0.050124298 | 0.053028591 |
| *DPB1*10.01* | 0.356531065 | -0.463065205 | 0.502242518 |
| *DPB1*04.01* | 0.363832621 | -0.030561811 | 0.033655183 |
| *DRB1*07* | 0.369076509 | 0.909561811 | 1.012644406 |
| *C*14.02* | 0.371162235 | -0.127857839 | 0.14296999 |
| *DRB1*01.01* | 0.375502581 | 0.08546352 | 0.096436734 |
| *DRB1*04.08* | 0.386272365 | 0.123456228 | 0.142493795 |
| *DPB1*11.01* | 0.393085415 | 0.287013269 | 0.336067209 |
| *DRB4*ng* | 0.394047381 | -0.032487863 | 0.038117854 |
| *DRB4*01.01* | 0.398060478 | 0.144142756 | 0.170564935 |
| *A*03* | 0.399085649 | 0.378475106 | 0.448826214 |
| *DQB1*06.09* | 0.409607058 | 0.263277831 | 0.31928513 |
| *A*11.01* | 0.416308895 | -0.068236787 | 0.083948446 |
| *DQA1*02.01* | 0.422512168 | -0.058888893 | 0.073421092 |
| *B*13.01* | 0.425290813 | 0.002872386 | 0.00360275 |
| *B*46.01* | 0.425290813 | 0.002872386 | 0.00360275 |
| *C*04.06* | 0.425290813 | 0.002872386 | 0.00360275 |
| *DRB1*07.03* | 0.425290813 | 0.002872386 | 0.00360275 |
| *DRB5*01.02* | 0.425290813 | 0.002872386 | 0.00360275 |
| *B*44.03* | 0.428309855 | 0.131755228 | 0.166339304 |
| *DRB1*04.03* | 0.429181868 | -0.793309762 | 1.003437087 |
| *DRB4*01.03* | 0.433293001 | 0.031026016 | 0.039595763 |
| *DPB1*03.01* | 0.435011294 | -0.037909704 | 0.048562026 |
| *A*23.01* | 0.435618526 | 0.191579959 | 0.245737161 |
| *DRB1*07.01* | 0.441215485 | -0.056564546 | 0.073446876 |
| *DRB1*13.01* | 0.45099011 | 0.040577874 | 0.053833559 |
| *B*08.01* | 0.471818645 | -0.041365541 | 0.057490216 |
| *DQA1*04.01* | 0.482971672 | 0.038179609 | 0.054423265 |
| *C*02.02* | 0.48450041 | 0.047487252 | 0.067927956 |
| *DPB1*16.01* | 0.495519549 | 0.143006142 | 0.20982293 |
| *DQB1*04.02* | 0.507941227 | 0.036032391 | 0.054425746 |
| *C*12.02* | 0.509829515 | -0.654690387 | 0.993304854 |
| *DRB1*01.02* | 0.510606175 | 0.20913135 | 0.317879954 |
| *B*39.06* | 0.511515131 | 0.33194621 | 0.505645339 |
| *C*04.01* | 0.514056745 | -0.033894526 | 0.051942928 |
| *A*30* | 0.518952307 | -0.289330229 | 0.44860096 |

| | | | |
|---|---|---|---|
| *B*07* | 0.518969816 | 0.323665363 | 0.501857948 |
| *DQB1*02.02* | 0.522488636 | -0.05335569 | 0.083431627 |
| *DRB1*08.01* | 0.523298238 | 0.034951354 | 0.054759552 |
| *B*35.02* | 0.525146846 | -0.448481473 | 0.70579012 |
| *DRB1*11.03* | 0.532798203 | -0.441815574 | 0.708332665 |
| *DQB1*02* | 0.54406888 | -0.609634392 | 1.004883702 |
| *DRB1*04.05* | 0.565088692 | 0.411517963 | 0.715310706 |
| *DQA1*01.04* | 0.573104777 | 0.080759344 | 0.143321235 |
| *B*44.27* | 0.574685902 | 0.102177837 | 0.182082029 |
| *B*27.02* | 0.574853444 | 0.231319613 | 0.412394711 |
| *DQB1*06.03* | 0.575428777 | 0.031407595 | 0.056077554 |
| *B*15.17* | 0.594921001 | -0.378093994 | 0.711079301 |
| *B*52.01* | 0.598430868 | -0.523589337 | 0.994172679 |
| *DPB1*01.01* | 0.598676443 | -0.037584768 | 0.071412549 |
| *DRB1*14.01* | 0.624633167 | 0.148384266 | 0.303261626 |
| *C*03.03* | 0.624675665 | -0.028822459 | 0.058913378 |
| *DQA1*03* | 0.625244236 | 0.113105372 | 0.231568547 |
| *DRB1*11.04* | 0.647473221 | 0.119005168 | 0.260248438 |
| *B*47.01* | 0.649063931 | -0.0900813 | 0.197953478 |
| *DQB1*05.02* | 0.658485585 | 0.069309128 | 0.156806207 |
| *B*58.01* | 0.666063305 | 0.116435094 | 0.269802257 |
| *C*12.03* | 0.671053406 | 0.038312853 | 0.090210991 |
| *DRB1*08.02* | 0.677688327 | 0.170863031 | 0.411104288 |
| *DQA1*01.02* | 0.683003911 | -0.01749075 | 0.042830909 |
| *B*51.01* | 0.683326192 | 0.027845215 | 0.068260044 |
| *DQB1*05.03* | 0.683340635 | 0.060941117 | 0.14939888 |
| *DPB1*19.01* | 0.695144405 | -0.095589037 | 0.243922168 |
| *DRB1*14.02* | 0.698261861 | -0.104660787 | 0.269975421 |
| *DPB1*105.01* | 0.699024632 | -0.174006109 | 0.450049811 |
| *DRB1*11.01* | 0.703623242 | -0.032982233 | 0.086696323 |
| *DPB1*05.01* | 0.706199433 | -0.041200281 | 0.10929464 |
| *A*32.01* | 0.70797917 | -0.03236834 | 0.086414317 |
| *A*30.01* | 0.710348179 | -0.124849098 | 0.336168442 |
| *DQA1*05.03* | 0.713074076 | -0.099283384 | 0.269989409 |
| *DRB3*01.01* | 0.713102623 | -0.015894322 | 0.043227228 |
| *C*07.01* | 0.715228902 | -0.018036774 | 0.049437137 |
| *DQB1*05.01* | 0.730772055 | 0.034050596 | 0.098955896 |
| *C*07.04* | 0.735296725 | 0.039176452 | 0.115876425 |
| *B*14.01* | 0.740724912 | -0.23539282 | 0.711385388 |
| *B*07.02* | 0.743718959 | -0.015587383 | 0.047677685 |
| *DQA1*01.03* | 0.746424658 | 0.018190684 | 0.056255675 |
| *DRB4*01.03N* | 0.747905234 | -0.050845882 | 0.158199919 |
| *DQA1*03.02* | 0.751024817 | 0.026052489 | 0.082109819 |
| *C*07.02* | 0.752375879 | -0.014532264 | 0.046059871 |
| *DPB1*17.01* | 0.758118665 | -0.080304472 | 0.260769231 |

| | | | |
|---|---|---|---|
| *A*02.01* | 0.75977546 | 0.010595326 | 0.034650652 |
| *A*25.01* | 0.763271679 | 0.045065707 | 0.149626852 |
| *DQB1*03.03* | 0.763331797 | 0.022381207 | 0.074329388 |
| *DPB1*20.01* | 0.770380768 | -0.065954248 | 0.225965997 |
| *C*07* | 0.771525613 | 0.288150841 | 0.992323617 |
| *DRB1*04.01* | 0.776546093 | 0.01596278 | 0.056242043 |
| *B*44.02* | 0.784397617 | 0.0180344 | 0.065916955 |
| *B*18* | 0.789567154 | -0.189739045 | 0.710972024 |
| *B*57.01* | 0.79739274 | 0.03955714 | 0.154085004 |
| *DPB1*15.01* | 0.801881865 | 0.057967017 | 0.231024952 |
| *C*03.02* | 0.805470127 | 0.066440214 | 0.269781671 |
| *DRB5*02.02* | 0.806355031 | 0.043869083 | 0.17896182 |
| *C*17.03* | 0.823948789 | 0.074699521 | 0.335774938 |
| *C*05.01* | 0.828364079 | 0.01508147 | 0.069563919 |
| *B*56.01* | 0.832953397 | -0.024637369 | 0.116811694 |
| *DRB3*ng* | 0.833420485 | -0.007748519 | 0.036842145 |
| *B*39.01* | 0.836796019 | -0.017819987 | 0.086507538 |
| *B*18.01* | 0.839350417 | 0.014958072 | 0.073785244 |
| *B*41.02* | 0.858972326 | 0.059660067 | 0.335767818 |
| *DRB1*13.03* | 0.863627084 | 0.03977395 | 0.231568508 |
| *B*15.01* | 0.86974395 | -0.007914025 | 0.048261038 |
| *B*35.01* | 0.877316345 | -0.00905613 | 0.058664291 |
| *DPB1*13.01* | 0.882382093 | -0.044948546 | 0.303808677 |
| *C*16.01* | 0.885178308 | -0.038859039 | 0.269092027 |
| *DRB1*14* | 0.889511811 | 0.139954885 | 1.007435228 |
| *DRB1*09.01* | 0.89357991 | -0.012368501 | 0.09245698 |
| *DPB1*06.01* | 0.896205651 | -10.46505812 | 80.21905385 |
| *B*39* | 0.8978483 | -10.35022304 | 80.62180499 |
| *B*50.01* | 0.900419576 | -0.062914559 | 0.502788195 |
| *DPB1*02.01* | 0.900557755 | -0.005887081 | 0.047112935 |
| *A*33.01* | 0.902931228 | -11.38615027 | 93.3602233 |
| *DRB1*01.03* | 0.905777298 | -10.2691666 | 86.75740043 |
| *C*04* | 0.913341438 | -8.493300837 | 78.04562324 |
| *DRB1*15.02* | 0.914338836 | -10.24494616 | 95.2420859 |
| *B*49.01* | 0.915697519 | 0.04780854 | 0.451642423 |
| *DQB1*06.01* | 0.917282449 | -10.25918993 | 98.78142379 |
| *A*02.02* | 0.919658178 | -9.348285764 | 92.68180307 |
| *A*02* | 0.921376295 | -10.35533107 | 104.9170021 |
| *B*15* | 0.922818602 | -10.37286445 | 107.0649045 |
| *A*29.01* | 0.924872881 | -0.031664658 | 0.335795508 |
| *C*15.05* | 0.926143008 | -0.093323113 | 1.006737 |
| *B*13* | 0.927462226 | -7.99337608 | 87.80242489 |
| *DRB1*04.07* | 0.92875121 | -0.040254626 | 0.450194262 |
| *B*15.16* | 0.93065132 | -7.177055707 | 82.47081302 |
| *DRB1*14.54* | 0.930989689 | 0.01459089 | 0.168486575 |

| | | | |
|---|---|---|---|
| *DQB1*03.04* | 0.933352692 | -9.354287779 | 111.8567518 |
| *C*12* | 0.933499717 | -8.232260257 | 98.65791172 |
| *DQB1*06* | 0.934882507 | -7.142504265 | 87.41982336 |
| *A*01* | 0.934927206 | -7.539230498 | 92.33903348 |
| *DQB1*04* | 0.935452897 | -7.428088524 | 91.72038006 |
| *C*01* | 0.937519658 | -6.106946104 | 77.9069443 |
| *C*16.02* | 0.940768965 | -7.101765328 | 95.57792182 |
| *A*32* | 0.940881652 | -6.375303521 | 85.96480501 |
| *A*02.17* | 0.94151451 | -7.627870102 | 103.9694511 |
| *C*03* | 0.942377052 | -6.239473182 | 86.32060277 |
| *A*31* | 0.944029688 | -8.617584147 | 122.7470943 |
| *DRB1*15* | 0.944078086 | -6.929607845 | 98.78947231 |
| *C*02* | 0.945464977 | -5.686734469 | 83.13596496 |
| *A*28* | 0.946271069 | -6.463839582 | 95.91660742 |
| *DPB1*09.01* | 0.946705368 | -6.993057668 | 104.6165467 |
| *C*06* | 0.948519501 | -5.384809059 | 83.39998309 |
| *A*30.02* | 0.949107542 | -7.184650497 | 112.5634705 |
| *A*11* | 0.949290816 | -7.999456234 | 125.7827908 |
| *DQA1*03.03* | 0.949496319 | 0.006340545 | 0.100104432 |
| *A*01.02* | 0.950824469 | -7.250852648 | 117.5722564 |
| *A*02.06* | 0.951612427 | -6.595284312 | 108.6859178 |
| *DRB1*13* | 0.952427398 | -6.594242861 | 110.5326382 |
| *A*68* | 0.953136965 | -6.532484879 | 111.1573375 |
| *B*40* | 0.953522329 | -6.556445482 | 112.4911435 |
| *B*08* | 0.954725374 | -5.480877539 | 96.53883087 |
| *B*27* | 0.956261475 | -5.999197471 | 109.3833951 |
| *DRB1*04.02* | 0.957538714 | -4.338950593 | 81.49415693 |
| *A*68167* | 0.959860119 | -5.474648481 | 108.7769576 |
| *C*05* | 0.963645546 | -5.445276344 | 119.4681128 |
| *DPB1*34.01* | 0.963647702 | -5.444951432 | 119.4680757 |
| *B*38.01* | 0.969208361 | -0.007269341 | 0.188319126 |
| *C*17.01* | 0.975270042 | 0.01107589 | 0.357294025 |
| *DRB1*16.01* | 0.976100131 | 0.005277518 | 0.176160813 |
| *B*51* | 0.984341139 | -0.008831391 | 0.449967496 |
| *B*44* | 0.989903146 | 0.009026446 | 0.713278584 |
| *B*45.01* | 0.991009604 | -0.005677486 | 0.503857944 |
| *A*24* | 0.991540039 | -0.007490415 | 0.706430478 |
| *A*31.01* | 0.992478594 | 0.000771807 | 0.081873486 |
| *C*15.02* | 0.992694124 | 0.000970448 | 0.105982501 |
| *B*41.01* | 0.995569611 | -0.001983862 | 0.357278945 |
| **SNP (rs9391858)** | **0.425290813** | **0.002872386** | **0.00360275** |

#### Table S4 – HLA fine mapping, respiratory tuberculosis

HLA fine mapping results for respiratory tuberculosis (ICD-10 A15-A16).

| HLA allele | P-value | Beta (Effect Estimate) | Standard Error |
|---|---|---|---|
| *DQB1*05.01* | 2.57E-05 | -0.217530205 | 0.051687048 |
| *DRB1*01.01* | 8.55E-05 | -0.207741553 | 0.052882634 |
| *DQA1*01.01* | 0.000156877 | -0.198018798 | 0.052386994 |
| *DRB1*13.02* | 0.000189008 | 0.333265549 | 0.089269005 |
| *DRB3*03.01* | 0.000428406 | 0.320379008 | 0.090966707 |
| *DQB1*06.04* | 0.000448685 | 0.324214769 | 0.092377899 |
| *DQB1*03.02* | 0.002020055 | 0.172745091 | 0.055954034 |
| *DQA1*03.01* | 0.002520448 | 0.169633305 | 0.056153686 |
| *DRB1*04.04* | 0.005310594 | 0.236456056 | 0.084825333 |
| *A*69.01* | 0.006040183 | 1.140837499 | 0.415516035 |
| *B*40.01* | 0.008854006 | 0.192982289 | 0.073723759 |
| *C*03.04* | 0.010381416 | 0.140631428 | 0.054872867 |
| *C*04.01* | 0.011311087 | -0.138904535 | 0.054839289 |
| *DQA1*04.01* | 0.019412036 | 0.143601615 | 0.061433257 |
| *DRB1*08.01* | 0.025257602 | 0.138440248 | 0.061874398 |
| *DQB1*04.02* | 0.025443048 | 0.137489485 | 0.06152732 |
| *C*08* | 0.036941869 | 1.22512111 | 0.587192117 |
| *DQA1*01.05* | 0.040698176 | -0.550208599 | 0.268841391 |
| *A*01.01* | 0.045676688 | -0.142239647 | 0.071177875 |
| *DPB1*14.01* | 0.055112684 | 0.277777522 | 0.14482765 |
| *DRB1*10.01* | 0.055935898 | -0.496455807 | 0.259715819 |
| *DRB3*ng* | 0.057247981 | -0.077082693 | 0.040539697 |
| *B*35.01* | 0.057875839 | -0.112208471 | 0.059161801 |
| *B*55.01* | 0.060648031 | -0.711656977 | 0.379337393 |
| *B*13.02* | 0.061103285 | -0.231111005 | 0.123407468 |
| *DRB1*13.01* | 0.083443599 | 0.107176186 | 0.061914126 |
| *C*06.02* | 0.102287668 | -0.138030101 | 0.084480841 |
| *DRB4*01.03* | 0.10760274 | 0.073428054 | 0.045634068 |
| *B*15.01* | 0.110784972 | 0.087442852 | 0.054834284 |
| *A*03* | 0.133533435 | 0.673322682 | 0.448788246 |
| *DQB1*06.03* | 0.137654214 | 0.09594982 | 0.064630805 |
| *DPB1*19.01* | 0.143817717 | -0.518095084 | 0.354441875 |
| *A*24.02* | 0.153392896 | 0.091697695 | 0.064229857 |
| *C*08.02* | 0.155878573 | -0.709555326 | 0.50001428 |
| *B*35.08* | 0.160607655 | 0.447732816 | 0.319118309 |
| *DRB4*ng* | 0.190742766 | -0.058002702 | 0.044331525 |
| *DQA1*01.03* | 0.196645105 | 0.08369952 | 0.064824608 |
| *DRB1*01.02* | 0.197395612 | 0.431266274 | 0.334572453 |
| *DRB1*09.01* | 0.197597044 | -0.142966476 | 0.110961951 |
| *DQB1*06.02* | 0.202260153 | -0.072699444 | 0.05701297 |
| *DRB1*15.01* | 0.209818097 | -0.071147768 | 0.056733681 |

| | | | |
|---|---|---|---|
| *DRB5*01.01* | 0.210705893 | -0.071085143 | 0.056794201 |
| *DRB5*ng* | 0.222977391 | 0.06756268 | 0.055440634 |
| *A*33.03* | 0.238517625 | 0.397286827 | 0.337055062 |
| *DQA1*02.01* | 0.23904303 | -0.102354823 | 0.086934244 |
| *DRB1*12.01* | 0.239172278 | -0.15030753 | 0.127697619 |
| *DRB4*01.03N* | 0.239527844 | -0.209971625 | 0.178521833 |
| *B*37.01* | 0.242785215 | -0.293990547 | 0.251692722 |
| *DRB1*07.01* | 0.249712458 | -0.10009863 | 0.086963031 |
| *DRB1*04.01* | 0.252177962 | 0.07417319 | 0.064775818 |
| *DQB1*03.03* | 0.276869159 | -0.094193077 | 0.086623928 |
| *DRB1*04.05* | 0.278332176 | 0.775989469 | 0.715808856 |
| *C*02.02* | 0.282036589 | 0.084255642 | 0.078322292 |
| *DRB1*07* | 0.29469067 | 1.062018698 | 1.013485894 |
| *B*13.01* | 0.321408472 | 0.004186826 | 0.004222431 |
| *B*46.01* | 0.321408472 | 0.004186826 | 0.004222431 |
| *C*04.06* | 0.321408472 | 0.004186826 | 0.004222431 |
| *DRB1*07.03* | 0.321408472 | 0.004186826 | 0.004222431 |
| *DRB5*01.02* | 0.321408472 | 0.004186826 | 0.004222431 |
| *B*39.24* | 0.323889227 | -0.989250758 | 1.002791326 |
| *DPB1*01.01* | 0.325233809 | -0.084732404 | 0.086131223 |
| *C*05.01* | 0.327624735 | 0.078588391 | 0.080281576 |
| *DQA1*01.04* | 0.349420495 | 0.155437021 | 0.166115963 |
| *DPB1*105.01* | 0.356097069 | -0.534035979 | 0.578695299 |
| *DQA1*05.01* | 0.366897939 | -0.058852973 | 0.065225561 |
| *DRB3*01.01* | 0.371472631 | 0.044015957 | 0.04925037 |
| *DRB1*03.01* | 0.374904718 | -0.05797035 | 0.065331692 |
| *C*14.02* | 0.390438673 | -0.148483362 | 0.172891874 |
| *DQB1*02.01* | 0.391868847 | -0.055947598 | 0.065341549 |
| *B*44.02* | 0.393012102 | 0.065374717 | 0.076536166 |
| *B*51* | 0.401360676 | 0.377601147 | 0.449957133 |
| *DRB1*04.08* | 0.405832675 | 0.140471253 | 0.168988068 |
| *DRB1*08.02* | 0.406552333 | 0.373225014 | 0.449681675 |
| *DRB1*04.07* | 0.416748866 | 0.365882628 | 0.450553014 |
| *A*26.01* | 0.434014676 | -0.123491281 | 0.157848571 |
| *DRB1*11.04* | 0.436320886 | -0.295334667 | 0.379402254 |
| *DQA1*05.03* | 0.439363263 | 0.2166679 | 0.280197358 |
| *C*03.03* | 0.45145163 | 0.050567767 | 0.067155324 |
| *DRB1*14.02* | 0.452205086 | 0.210626248 | 0.280183516 |
| *B*39.06* | 0.457846207 | 0.432627632 | 0.582743499 |
| *DQB1*05.03* | 0.460534029 | 0.127887897 | 0.1732974 |
| *DPB1*10.01* | 0.461889721 | -0.426225685 | 0.579316642 |
| *A*30* | 0.476928284 | -0.411388124 | 0.578400208 |
| *DPB1*15.01* | 0.477302505 | -0.225433853 | 0.31722303 |
| *DRB1*11.03* | 0.479719654 | -0.707001583 | 1.000350967 |
| *B*38.01* | 0.480527618 | -0.172836478 | 0.245000279 |

| | | | |
|---|---|---|---|
| *DQA1*05.05* | 0.488202626 | -0.051983204 | 0.074993399 |
| *B*35* | 0.491415305 | -0.688427916 | 1.000535422 |
| *DQB1*06.09* | 0.497150707 | 0.258264092 | 0.380370845 |
| *DPB1*17.01* | 0.511004332 | -0.220578851 | 0.335596162 |
| *B*08.01* | 0.517509745 | -0.043965782 | 0.067933469 |
| *C*01.02* | 0.517580244 | 0.047510838 | 0.073423451 |
| *B*40.02* | 0.522330504 | -0.078835203 | 0.123226766 |
| *A*02.05* | 0.526061767 | 0.369143377 | 0.582218259 |
| *C*07* | 0.531852107 | 0.617182361 | 0.987205489 |
| *DQA1*01.02* | 0.533946431 | 0.030520188 | 0.049068374 |
| *A*23.01* | 0.538975666 | 0.179294758 | 0.291838604 |
| *DQA1*03.02* | 0.548075909 | -0.058624362 | 0.097602214 |
| *B*14.02* | 0.548947218 | -0.24491906 | 0.408649009 |
| *DPB1*03.01* | 0.565189082 | 0.032026382 | 0.055683418 |
| *B*14.01* | 0.571060166 | -0.568316015 | 1.003221918 |
| *A*24* | 0.571550696 | 0.39896936 | 0.705180435 |
| *B*50.01* | 0.578211959 | -0.394223742 | 0.709037567 |
| *B*47.01* | 0.590203586 | 0.11571472 | 0.214867045 |
| *DQB1*02.02* | 0.591916281 | -0.052981431 | 0.098834675 |
| *DRB1*13.03* | 0.593692005 | 0.138811482 | 0.260193386 |
| *DPB1*20.01* | 0.593991832 | 0.13055689 | 0.244919547 |
| *DPB1*11.01* | 0.594374999 | 0.21873136 | 0.410757835 |
| *B*18* | 0.596772535 | -0.530610798 | 1.002955026 |
| *DPB1*16.01* | 0.602476102 | 0.130671965 | 0.250885486 |
| *DRB1*14.01* | 0.60752147 | 0.182465593 | 0.355257801 |
| *DQA1*03* | 0.61244736 | 0.136412281 | 0.269279172 |
| *B*27.05* | 0.635876649 | 0.033157566 | 0.070030716 |
| *B*07.02* | 0.637686988 | -0.026542513 | 0.056361282 |
| *DRB1*11.01* | 0.640152847 | 0.046508252 | 0.099486063 |
| *C*07.01* | 0.649877531 | -0.026424763 | 0.058213 |
| *B*44.03* | 0.658219997 | 0.089285775 | 0.201834122 |
| *DQA1*03.03* | 0.660552163 | 0.051430766 | 0.117114094 |
| *DRB1*14* | 0.666720517 | 0.433863557 | 1.007454508 |
| *C*12.02* | 0.677460991 | -0.412336856 | 0.991360413 |
| *DRB1*04.03* | 0.679165535 | -0.415108633 | 1.003642469 |
| *DPB1*04.01* | 0.68979877 | 0.015545046 | 0.038947504 |
| *DRB4*01.01* | 0.691667063 | 0.08325686 | 0.209929833 |
| *A*29.02* | 0.692784851 | -0.149918747 | 0.379465583 |
| *C*07.04* | 0.697382785 | 0.053192287 | 0.136791789 |
| *C*15.02* | 0.703973006 | -0.046229365 | 0.121668125 |
| *DRB1*14.54* | 0.707669143 | 0.074132548 | 0.197692946 |
| *B*56.01* | 0.709489548 | 0.049731334 | 0.133491982 |
| *A*02.01* | 0.737250162 | 0.013569172 | 0.040444981 |
| *DQA1*06.01* | 0.74107897 | -0.234791095 | 0.710573364 |
| *DRB3*02.02* | 0.742687745 | -0.020741709 | 0.063179871 |

| | | | |
|---|---|---|---|
| *A*11.01* | 0.743930658 | -0.031565983 | 0.096634724 |
| *A*30.01* | 0.747329636 | 0.114861888 | 0.356534075 |
| *B*27.02* | 0.747410639 | 0.162235456 | 0.50374997 |
| *DRB1*16.01* | 0.754311628 | -0.068831536 | 0.219937701 |
| *B*58.01* | 0.766768972 | 0.094506296 | 0.318628017 |
| *C*16.01* | 0.770321245 | 0.088508658 | 0.303158852 |
| *A*32.01* | 0.777058869 | -0.028704196 | 0.101373129 |
| *B*44* | 0.783174021 | 0.196574081 | 0.714333987 |
| *B*52.01* | 0.789153023 | -0.265281619 | 0.992038455 |
| *B*49.01* | 0.792688014 | -0.152761248 | 0.58123531 |
| *B*18.01* | 0.809531881 | 0.020943175 | 0.086890333 |
| *DPB1*13.01* | 0.818125205 | -0.081826135 | 0.355832373 |
| *DPB1*02.01* | 0.839983315 | -0.011186202 | 0.055400598 |
| *B*44.27* | 0.844672166 | 0.044231931 | 0.22576436 |
| *A*25.01* | 0.851320991 | 0.032576343 | 0.173802412 |
| *B*41.01* | 0.861475369 | -0.071678306 | 0.410772432 |
| *B*57.01* | 0.865205629 | 0.02861938 | 0.168595818 |
| *DPB1*05.01* | 0.869855445 | -0.020994069 | 0.12813598 |
| *DQB1*03.01* | 0.870416949 | -0.010017358 | 0.061407622 |
| *B*39.01* | 0.874091606 | 0.016105056 | 0.101632611 |
| *C*07.02* | 0.87557646 | -0.008470949 | 0.054100074 |
| *C*03.02* | 0.88812853 | 0.044824401 | 0.318643263 |
| *A*68.01* | 0.890343021 | 0.01098518 | 0.079677711 |
| *C*17.01* | 0.890665582 | -0.056467689 | 0.410787461 |
| *C*12.03* | 0.893996024 | -0.014484049 | 0.108698671 |
| *B*35.02* | 0.897583278 | -0.090750352 | 0.70504954 |
| *A*33.01* | 0.906115418 | -11.02764303 | 93.50236442 |
| *DPB1*02.02* | 0.907730493 | -11.04115854 | 95.26317489 |
| *DQB1*02* | 0.909627699 | -10.24297187 | 90.23996525 |
| *A*03.01* | 0.916713888 | 0.004624962 | 0.044226713 |
| *DRB1*15.02* | 0.91674152 | -9.944333318 | 95.12546752 |
| *DPB1*04.02* | 0.917432499 | -0.005033157 | 0.048550538 |
| *DQB1*06.01* | 0.919640884 | -9.952211074 | 98.64799944 |
| *A*02.02* | 0.922921829 | -8.982238713 | 92.83592731 |
| *C*15.05* | 0.922929424 | -10.08147524 | 104.2073866 |
| *A*02* | 0.92302514 | -10.12312672 | 104.7684395 |
| *B*15* | 0.925328118 | -10.02786094 | 106.99311 |
| *B*13* | 0.93090064 | -7.627581647 | 87.96480317 |
| *DRB1*08.03* | 0.932879731 | 0.048957505 | 0.581289463 |
| *B*45.01* | 0.933033469 | 0.048815561 | 0.580937887 |
| *DPB1*06.01* | 0.933402199 | -11.06227817 | 132.3791314 |
| *B*39* | 0.933747343 | -11.03915689 | 132.7922343 |
| *A*29.01* | 0.934995692 | -0.03096161 | 0.3796122 |
| *C*12* | 0.935542166 | -7.964395469 | 98.47907356 |
| *DQB1*03.04* | 0.935757669 | -9.026491713 | 111.9870248 |

| | | | |
|---|---|---|---|
| *DRB1*01.03* | 0.937791431 | -11.15696027 | 142.9535762 |
| *DQB1*04* | 0.938268392 | -7.047019873 | 90.99216474 |
| *A*01* | 0.938499553 | -7.151345856 | 92.68700471 |
| *DQB1*05.02* | 0.941150966 | 0.014365208 | 0.19458901 |
| *C*16.02* | 0.942290252 | -6.826854708 | 94.30448617 |
| *C*04* | 0.942824693 | -9.21623848 | 128.5029409 |
| *A*02.17* | 0.943018931 | -7.442331472 | 104.1235131 |
| *DRB1*15* | 0.945735197 | -6.721509188 | 98.75370748 |
| *B*07* | 0.946338149 | 0.047643494 | 0.707864639 |
| *A*31* | 0.946856862 | -8.210411495 | 123.1788953 |
| *DPB1*09.01* | 0.948387374 | -6.7890517 | 104.8793544 |
| *A*28* | 0.948769897 | -6.135350941 | 95.48947649 |
| *A*11* | 0.950232772 | -7.792465451 | 124.8503028 |
| *B*41.02* | 0.950252611 | -0.025591254 | 0.410184896 |
| *A*30.02* | 0.951940519 | -6.834800084 | 113.4028405 |
| *A*02.06* | 0.952504957 | -6.481188573 | 108.8152372 |
| *A*01.02* | 0.953416224 | -6.937479408 | 118.7572475 |
| *A*68* | 0.953426993 | -6.482957609 | 111.0023378 |
| *B*15.16* | 0.954205101 | -7.851748893 | 136.7258734 |
| *B*40* | 0.954382022 | -6.367360245 | 111.3080657 |
| *DRB1*13* | 0.954452755 | -6.302121987 | 110.3389067 |
| *DQB1*06* | 0.955780047 | -7.849654531 | 141.5630196 |
| *C*01* | 0.957308714 | -6.883030184 | 128.5799118 |
| *B*08* | 0.957325599 | -5.135419793 | 95.97129603 |
| *B*27* | 0.957957241 | -5.740715729 | 108.8964633 |
| *C*03* | 0.961152192 | -6.942724754 | 142.5383709 |
| *A*32* | 0.961756875 | -6.787938563 | 141.5657511 |
| *A*68167* | 0.962227983 | -5.164163199 | 109.0454446 |
| *C*02* | 0.96308987 | -6.331952393 | 136.8286803 |
| *C*06* | 0.96432171 | -6.147534568 | 137.433371 |
| *B*35.03* | 0.965295534 | -0.030943522 | 0.711192935 |
| *DPB1*34.01* | 0.965994633 | -5.093201337 | 119.4680853 |
| *C*05* | 0.966377526 | -5.035821305 | 119.4681419 |
| *DRB1*04.02* | 0.970881375 | -4.922582852 | 134.8546107 |
| *A*31.01* | 0.975180418 | -0.003004782 | 0.096580277 |
| *B*15.17* | 0.977968601 | -0.019644059 | 0.711334754 |
| *C*17.03* | 0.981051403 | -0.009742009 | 0.410176429 |
| *DRB5*02.02* | 0.982737139 | 0.004761255 | 0.220046689 |
| *B*51.01* | 0.997623636 | 0.000238906 | 0.080214726 |

#### Table S5 – Epidemiological correlates

| Trait | TB N | TB % | CTRL N | CTRL % | FG R7 N | FG R7 % | P unadj. | P adj. | beta (adj.) | OR (adj.) | SE adj. | CI 95% low | CI 95% high |
|---|---|---|---|---|---|---|---|---|---|---|---|---|---|
| Ever smoker | 928 | 66.43% | 95404 | 49.63% | 96332 | 49.75% | 1.30E-34 | 2.00E-16 | 0.63 | 1.87 | 0.06 | 1.75 | 1.99 |
| Current smoker | 598 | 42.81% | 49898 | 25.96% | 50496 | 26.08% | 2.60E-44 | 2.00E-16 | 0.66 | 1.94 | 0.06 | 1.82 | 2.06 |
| COPD | 402 | 21.21% | 12853 | 4.18% | 13255 | 4.29% | 1.40E-233 | 2.00E-16 | 1.31 | 3.71 | 0.07 | 3.57 | 3.85 |
| Diabetes | 377 | 19.89% | 53011 | 17.25% | 53388 | 17.27% | 2.50E-03 | 9.40E-02 | -0.11 | 0.89 | 0.07 | 0.75 | 1.03 |
| Hypertension | 324 | 17.10% | 84667 | 27.56% | 85491 | 27.65% | 8.20E-52 | 5.60E-02 | 0.11 | 1.11 | 0.06 | 0.99 | 1.23 |
| Asthma | 377 | 19.89% | 31974 | 10.41% | 32351 | 10.46% | 2.80E-163 | 2.00E-16 | 2.40 | 10.97 | 0.11 | 10.75 | 11.19 |
| Major cardiac event (CHD) | 784 | 41.37% | 80123 | 26.08% | 80907 | 26.17% | 1.30E-49 | 3.80E-03 | 0.16 | 1.17 | 0.05 | 1.07 | 1.27 |
| HIV | 9 | 0.47% | 418 | 0.14% | 427 | 0.14% | 2.10E-04 | 1.30E-01 | 0.87 | 2.40 | 0.58 | 1.26 | 3.54 |
| Rheumatoid arthritis | 122 | 6.44% | 9733 | 3.17% | 9855 | 3.19% | 1.20E-18 | 1.10E-05 | 0.50 | 1.65 | 0.11 | 1.43 | 1.87 |
| Biological medication for rheumatoid arthritis | 23 | 1.21% | 3116 | 1.01% | 3139 | 1.02% | 3.90E-01 | 1.10E-02 | 0.65 | 1.91 | 0.25 | 1.42 | 2.40 |
| Sleep apnea | 175 | 9.23% | 27032 | 8.80% | 27207 | 8.80% | 5.10E-01 | 6.90E-01 | 0.04 | 1.10 | 0.09 | 0.92 | 1.28 |
| Alcohol dependence | 123 | 6.49% | 8014 | 2.61% | 8137 | 2.63% | 5.20E-25 | 2.00E-16 | 0.91 | 2.50 | 0.10 | 2.30 | 2.70 |
| Alcohol use disorder | 157 | 8.28% | 12505 | 4.07% | 12662 | 4.10% | 2.00E-19 | 1.10E-15 | 0.75 | 2.11 | 0.09 | 1.93 | 2.29 |
| Inflammatory bowel disease | 74 | 3.91% | 8630 | 2.81% | 8704 | 2.82% | 4.20E-03 | 4.50E-05 | 0.53 | 1.71 | 0.13 | 1.46 | 1.96 |
| Crohn's disease | 12 | 0.63% | 1533 | 0.50% | 1545 | 0.50% | 3.70E-01 | 2.00E-02 | 0.71 | 2.04 | 0.31 | 1.43 | 2.65 |
| **Tuberculosis all organs** | | | | | **1895** | **0.61%** | | | | | | | |

Trait = Studied risk factor or comorbidity
TB = Tuberculosis cases (ICD-10 A15-A19, all organs)
CTRL = Controls
FG = FinnGen data freeze R7
N = Number of individuals
% = Percentage of individuals with the studied trait (within TB cases, controls or FinnGen data freeze R7)
Adj. = Value comes from the adjusted multivariate logistic regression model (adjusted with age at death or end of follow up (12/31/2019), BMI, sex and the first 10 genetic principal components)

P = P-Value
Beta = Effect estimate
OR = Odds ratio
SE = Standard error
CI 95% = 95 % confidence intervals for odds ratio

Large Registry Based Analysis of Genetic Predisposition to Tuberculosis
Identifies Genetic Risk Factors at HLA
Supplements

#### Table S6 – Cox proportional hazards model

| Trait | P-value unadj. | P-value adj. | SE | Coef. | HR | CI 95% low | CI 95% high | Global P-value of trait | N cases | N controls |
|---|---|---|---|---|---|---|---|---|---|---|
| Current smoker | 2.00E-16 | 3.27E-10 | 0.176 | 0.900 | 2.461 | 1.742 | 3.476 | 0.039 | 596 | 798 |
| Former smoker | 2.61E-02 | 3.31E-01 | 0.168 | 0.163 | 1.177 | 0.847 | 1.635 | 0.039 | 330 | 1064 |
| COPD | 2.00E-16 | 8.98E-10 | 0.090 | 0.548 | 1.730 | 1.452 | 2.062 | 0.002 | 402 | 992 |
| Asthma | 9.76E-01 | 3.47E-01 | 0.108 | 0.101 | 1.107 | 0.896 | 1.367 | 0.037 | 377 | 1017 |
| Major cardiac event (CHD) | 2.90E-01 | 8.55E-01 | 0.093 | -0.017 | 0.983 | 0.819 | 1.180 | 0.046 | 784 | 610 |
| Rheumatoid arthritis | 2.64E-01 | 4.43E-01 | 0.223 | -0.171 | 0.843 | 0.544 | 1.310 | 0.068 | 78 | 1316 |
| Alcohol dependence | 2.67E-09 | 4.20E-07 | 0.160 | 0.807 | 2.240 | 1.640 | 3.060 | 0.090 | 100 | 1294 |
| Alcohol use disorder | 9.48E-09 | 1.45E-06 | 0.147 | 0.708 | 2.030 | 1.520 | 2.710 | 0.091 | 122 | 1272 |
| Inflammatory bowel disease | 8.22E-01 | 5.83E-01 | 0.271 | -0.149 | 0.862 | 0.506 | 1.470 | 0.048 | 48 | 1346 |

Trait = Studied risk factor or comorbidity
N = Number of individuals
Adj. = Value comes from the adjusted Cox proportional hazards model (adjusted with stratified sex, stratified cohort (cohort representing for example biobank or study included within the FinnGen study), BMI and the first 10 genetic principal components)
P = P-Value
Coef. = Regression coefficient
HR = Hazard ratio
SE = Standard error
CI 95% = 95 % confidence intervals for hazard ratio

#### Table S7 – Changes of smoking over the decades

Description of smoking status among tuberculosis cases and matched controls throughout different decades starting from 1970s (time of the first diagnoses of tuberculosis in FinnGen data freeze R7).

| **Decade** | **TB ever smoker (%)** | **Matched CTRL ever smoker (%)** |
|---|---:|---:|
| 1970 - 1979 | 64.30 % | 57.00 % |
| 1980 - 1989 | 66.70 % | 62.70 % |
| 1990 - 1999 | 74.40 % | 60.20 % |
| 2000 - 2009 | 63.40 % | 55.90 % |
| 2010 - 2019 | 67.40 % | 52.20 % |
| **Average** | **67.24 %** | **57.60 %** |

TB = Tuberculosis (A15-A19, all organs)
CTRL = Control
Ever smoker = Smoking status "current" or "former"
% = Proportion of individuals who were ever smokers among cases or controls

*NB! Controls were matched according to age and sex with the tuberculosis cases using R programming language.

### Table S8 – Genetic Correlation TB vs. AUD

Genetic correlation results for tuberculosis (TB) and alcohol use disorder (AUD). Genetic correlation was conducted using three different AUD traits: Alcohol Use Disorders Identification Test – Consumption (AUDIT-C), Alcohol Use Disorders Identification Test – Problematic consequences of drinking (AUDIT-P) and Alcohol Use Disorders Identification Test – Total score (AUDIT-T).

| Trait 1 | Trait 2 | h2 | Genetic Correlation | Z-score | P-value |
|---|---|---|---|---|---|
| TB | AUDIT-C | 0.0839 (0.0055) | 0.1 (0.1192) | 0.8387 | 0.4016 |
| TB | AUDIT-P | 0.0584 (0.0048) | 0.1378 (0.1431) | 0.963 | 0.3355 |
| TB | AUDIT-T | 0.0919 (0.0059) | 0.1024 (0.1176) | 0.8708 | 0.3839 |

h2 = heritability of the trait 2

### Supplemental Information

#### Previous HLA findings from the literature

Our results align with previous studies showing the association between HLA class II region and tuberculosis:

An Icelandic GWAS study using cohort from Iceland, Croatia and Russia, showed susceptible association between tuberculosis and *HLA DQA1*03:01* allele (Sveinbjornsson et al., 2016). In addition, different HLA class II alleles have been associated with tuberculosis in, for example, different Asian and African populations (reviewed in Oliveira-Cortez et al., 2016). GWAS and HLA fine mapping in the Han Chinese population identified *HLA DQB1*02:01* having a significant association with tuberculosis (Qi et al., 2017). Additionally, HLA class II alleles *HLA DRB1*04:01* (Seedat et al., 2021 (South African)), *HLA DRB1*15:01* (Seedat et al., 2021 (South African)), *HLA DRB1*11:01* (Seedat et al., 2021 (South African)), *HLA DRB1*09:01* (Toyo-Oka et al., 2017 (Southeast Asian)), *HLA DQB1*03:03* (Toyo-Oka et al., 2017 (Southeast Asian)), *HLA DRB1*01:01* (Kim et al., 2005 (Korean); Amirzargar et al., 2004 (Iranian)), *HLA DQA1*01:01* (Amirzargar et al., 2004 (Iranian); Terán-Escandón et al., 1999 (Mexican)), *HLA DQB1*05:03* (Goldfeld et al., 1998 (Cambodian)), *HLA DQA1*05:05* (Li et al., 2021 (Chinese)) and gene *HLA DRA* (Bhattacharyya et al., 2019 (Indian)) have been associated with tuberculosis. In addition, *HLA DQA1*03* was associated with susceptibility of young onset tuberculosis in a Chinese cohort (Tang et al., 2019). Our finding of association between tuberculosis and *HLA DQB1*05:01* have previously been shown in South Indian (Ravikumar et al., 1999) and Mexican (Terán-Escandón et al., 1999) cohorts, but not in Caucasians nor in an as large cohort as FinnGen is. Furthermore, Tian et al. (2017) showed HLA class II genome-wide association among individuals with self-reported positivity in tuberculosis skin test using the 23andMe-study and approximately 100 000 individuals of European ancestry.

### Contributors of FinnGen

#### Steering Committee


| | |
|---|---|
| Aarno Palotie | Institute for Molecular Medicine Finland, HiLIFE, University of Helsinki, Finland |
| Mark Daly | Institute for Molecular Medicine Finland, HiLIFE, University of Helsinki, Finland |

Pharmaceutical companies

| | |
|---|---|
| Bridget Riley-Gills | Abbvie, Chicago, IL, United States |
| Howard Jacob | Abbvie, Chicago, IL, United States |
| Dirk Paul | Astra Zeneca, Cambridge, United Kingdom |
| Heiko Runz | Biogen, Cambridge, MA, United States |
| Sally John | Biogen, Cambridge, MA, United States |
| George Okafo | Boehringer Ingelheim, Ingelheim am Rhein, Germany |
| Nathan Lawless | Boehringer Ingelheim, Ingelheim am Rhein, Germany |
| Robert Plenge | Celgene, Summit, NJ, United States/Bristol Myers Squibb, New York, NY, United States |
| Joseph Maranville | Celgene, Summit, NJ, United States/Bristol Myers Squibb, New York, NY, United States |
| Mark McCarthy | Genentech, San Francisco, CA, United States |
| Julie Hunkapiller | Genentech, San Francisco, CA, United States |
| Meg Ehm | GlaxoSmithKline, Brentford, United Kingdom |
| Kirsi Auro | GlaxoSmithKline, Brentford, United Kingdom |
| Simonne Longerich | Merck, Kenilworth, NJ, United States |
| Caroline Fox | Merck, Kenilworth, NJ, United States |
| Anders Mälarstig | Pfizer, New York, NY, United States |
| Katherine Klinger | Sanofi, Paris, France |
| Deepak Raipal | Sanofi, Paris, France |
| Eric Green | Maze Therapeutics, San Francisco, CA, United States |
| Robert Graham | Maze Therapeutics, San Francisco, CA, United States |
| Robert Yang | Janssen Biotech, Beerse, Belgium |

| | |
|---|---|
| Chris O´Donnell | Novartis, Basel, Switzerland |

University of Helsinki & Biobanks

| | |
|---|---|
| Tomi Mäkelä | HiLIFE, University of Helsinki, Finland, Finland |
| Jaakko Kaprio | Institute for Molecular Medicine Finland, HiLIFE, Helsinki, Finland, Finland |
| Petri Virolainen | Auria Biobank / University of Turku / Hospital District of Southwest Finland, Turku, Finland |
| Antti Hakanen | Auria Biobank / University of Turku / Hospital District of Southwest Finland, Turku, Finland |
| Terhi Kilpi | THL Biobank / The National Institute of Health and Welfare Helsinki, Finland |
| Markus Perola | THL Biobank / The National Institute of Health and Welfare Helsinki, Finland |
| Jukka Partanen | Finnish Red Cross Blood Service / Finnish Hematology Registry and Clinical Biobank, Helsinki, Finland |
| Anne Pitkäranta | Helsinki Biobank / Helsinki University and Hospital District of Helsinki and Uusimaa, Helsinki |
| Juhani Junttila | Northern Finland Biobank Borealis / University of Oulu / Northern Ostrobothnia Hospital District, Oulu, Finland |
| Raisa Serpi | Northern Finland Biobank Borealis / University of Oulu / Northern Ostrobothnia Hospital District, Oulu, Finland |
| Tarja Laitinen | Finnish Clinical Biobank Tampere / University of Tampere / Pirkanmaa Hospital District, Tampere, Finland |
| Veli-Matti Kosma | Biobank of Eastern Finland / University of Eastern Finland / Northern Savo Hospital District, Kuopio, Finland |
| Jari Laukkanen | Central Finland Biobank / University of Jyväskylä / Central Finland Health Care District, Jyväskylä, Finland |
| Marco Hautalahti | FINBB - Finnish biobank cooperative |

Other Experts/ Non-Voting Members

| | |
|---|---|
| Outi Tuovila | Business Finland, Helsinki, Finland |
| Raimo Pakkanen | Business Finland, Helsinki, Finland |

Large Registry Based Analysis of Genetic Predisposition to Tuberculosis
Identifies Genetic Risk Factors at HLA
Supplements

**Scientific Committee**

Pharmaceutical companies

| | |
|---|---|
| Jeffrey Waring | Abbvie, Chicago, IL, United States |
| Bridget Riley-Gillis | Abbvie, Chicago, IL, United States |
| Ioanna Tachmazidou | Astra Zeneca, Cambridge, United Kingdom |
| Chia-Yen Chen | Biogen, Cambridge, MA, United States |
| Heiko Runz | Biogen, Cambridge, MA, United States |
| Zhihao Ding | Boehringer Ingelheim, Ingelheim am Rhein, Germany |
| Marc Jung | Boehringer Ingelheim, Ingelheim am Rhein, Germany |
| Shameek Biswas | Celgene, Summit, NJ, United States/Bristol Myers Squibb, New York, NY, United States |
| Sarah Pendergrass | Genentech, San Francisco, CA, United States |
| Julie Hunkapiller | Genentech, San Francisco, CA, United States |
| Meg Ehm | GlaxoSmithKline, Brentford, United Kingdom |
| David Pulford | GlaxoSmithKline, Brentford, United Kingdom |
| Neha Raghavan | Merck, Kenilworth, NJ, United States |
| Adriana Huertas-Vazquez | Merck, Kenilworth, NJ, United States |
| Jae-Hoon Sul | Merck, Kenilworth, NJ, United States |
| Anders Mälarstig | Pfizer, New York, NY, United States |
| Xinli Hu | Pfizer, New York, NY, United States |
| Katherine Klinger | Sanofi, Paris, France |
| Matthias Gossel | Sanofi, Paris, France |
| Robert Graham | Maze Therapeutics, San Francisco, CA, United States |
| Eric Green | Maze Therapeutics, San Francisco, CA, United States |
| Sahar Mozaffari | Maze Therapeutics, San Francisco, CA, United States |
| Dawn Waterworth | Janssen Research & Development, LLC, Spring House, PA, United States |
| Nicole Renaud | Novartis, Basel, Switzerland |
| Ma´en Obeidat | Novartis, Basel, Switzerland |

Large Registry Based Analysis of Genetic Predisposition to Tuberculosis
Identifies Genetic Risk Factors at HLA
Supplements

University of Helsinki & Biobanks


| | |
|---|---|
| Samuli Ripatti | Institute for Molecular Medicine Finland, HiLIFE, Helsinki, Finland |
| Johanna Schleutker | Auria Biobank / Univ. of Turku / Hospital District of Southwest Finland, Turku, Finland |
| Markus Perola | THL Biobank / The National Institute of Health and Welfare Helsinki, Finland |
| Mikko Arvas | Finnish Red Cross Blood Service / Finnish Hematology Registry and Clinical Biobank, Helsinki, Finland |
| Olli Carpén | Helsinki Biobank / Helsinki University and Hospital District of Helsinki and Uusimaa, Helsinki |
| Reetta Hinttala | Northern Finland Biobank Borealis / University of Oulu / Northern Ostrobothnia Hospital District, Oulu, Finland |
| Johannes Kettunen | Northern Finland Biobank Borealis / University of Oulu / Northern Ostrobothnia Hospital District, Oulu, Finland |
| Arto Mannermaa | Biobank of Eastern Finland / University of Eastern Finland / Northern Savo Hospital District, Kuopio, Finland |
| Katriina Aalto-Setälä | Finnish Clinical Biobank Tampere / University of Tampere / Pirkanmaa Hospital District, Tampere, Finland |
| Mika Kähönen | Finnish Clinical Biobank Tampere / University of Tampere / Pirkanmaa Hospital District, Tampere, Finland |
| Jari Laukkanen | Central Finland Biobank / University of Jyväskylä / Central Finland Health Care District, Jyväskylä, Finland |
| Johanna Mäkelä | FINBB - Finnish biobank cooperative |


Clinical Groups

<u>Neurology Group</u>


| | |
|---|---|
| Reetta Kälviäinen | Northern Savo Hospital District, Kuopio, Finland |
| Valtteri Julkunen | Northern Savo Hospital District, Kuopio, Finland |
| Hilkka Soininen | Northern Savo Hospital District, Kuopio, Finland |
| Anne Remes | Northern Ostrobothnia Hospital District, Oulu, Finland |
| Mikko Hiltunen | Northern Savo Hospital District, Kuopio, Finland |
| Jukka Peltola | Pirkanmaa Hospital District, Tampere, Finland |
| Minna Raivio | Hospital District of Helsinki and Uusimaa, Helsinki, Finland |
| Pentti Tienari | Hospital District of Helsinki and Uusimaa, Helsinki, Finland |

Large Registry Based Analysis of Genetic Predisposition to Tuberculosis
Identifies Genetic Risk Factors at HLA
Supplements


| | |
|---|---|
| Juha Rinne | Hospital District of Southwest Finland, Turku, Finland |
| Roosa Kallionpää | Hospital District of Southwest Finland, Turku, Finland |
| Juulia Partanen | Institute for Molecular Medicine Finland, HiLIFE, University of Helsinki, Finland |
| Ali Abbasi | Abbvie, Chicago, IL, United States |
| Adam Ziemann | Abbvie, Chicago, IL, United States |
| Jeffrey Waring | Abbvie, Chicago, IL, United States |
| Nizar Smaoui | Abbvie, Chicago, IL, United States |
| Anne Lehtonen | Abbvie, Chicago, IL, United States |
| Susan Eaton | Biogen, Cambridge, MA, United States |
| Heiko Runz | Biogen, Cambridge, MA, United States |
| Sanni Lahdenperä | Biogen, Cambridge, MA, United States |
| Janet van Adelsberg | Celgene, Summit, NJ, United States/ Bristol Myers Squibb, New York, NY, United States |
| Shameek Biswas | Celgene, Summit, NJ, United States/ Bristol Myers Squibb, New York, NY, United States |
| Julie Hunkapiller | Genentech, San Francisco, CA, United States |
| Natalie Bowers | Genentech, San Francisco, CA, United States |
| Edmond Teng | Genentech, San Francisco, CA, United States |
| Sarah Pendergrass | Genentech, San Francisco, CA, United States |
| Fanli Xu | GlaxoSmithKline, Brentford, United Kingdom |
| David Pulford | GlaxoSmithKline, Brentford, United Kingdom |
| Kirsi Auro | GlaxoSmithKline, Brentford, United Kingdom |
| Laura Addis | GlaxoSmithKline, Brentford, United Kingdom |
| John Eicher | GlaxoSmithKline, Brentford, United Kingdom |
| Qingqin S Li | Janssen Research & Development, LLC, Titusville, NJ 08560, United States |
| Karen He | Janssen Research & Development, LLC, Spring House, PA, United States |
| Ekaterina Khramtsova | Janssen Research & Development, LLC, Spring House, PA, United States |
| Beryl Cummings | Maze Therapeutics, San Francisco, CA, United States |
| Neha Raghavan | Merck, Kenilworth, NJ, United States |
| Kari Linden | Pfizer, New York, NY, United States |

Large Registry Based Analysis of Genetic Predisposition to Tuberculosis
Identifies Genetic Risk Factors at HLA
Supplements


Gastroenterology Group

| | |
|---|---|
| Martti Färkkilä | Hospital District of Helsinki and Uusimaa, Helsinki, Finland |
| Jukka Koskela | Hospital District of Helsinki and Uusimaa, Helsinki, Finland |
| Sampsa Pikkarainen | Hospital District of Helsinki and Uusimaa, Helsinki, Finland |
| Airi Jussila | Pirkanmaa Hospital District, Tampere, Finland |
| Katri Kaukinen | Pirkanmaa Hospital District, Tampere, Finland |
| Timo Blomster | Northern Ostrobothnia Hospital District, Oulu, Finland |
| Mikko Kiviniemi | Northern Savo Hospital District, Kuopio, Finland |
| Markku Voutilainen | Hospital District of Southwest Finland, Turku, Finland |
| Mark Daly | Institute for Molecular Medicine Finland, HiLIFE, University of Helsinki, Finland |
| Ali Abbasi | Abbvie, Chicago, IL, United States |
| Graham Heap | Abbvie, Chicago, IL, United States |
| Jeffrey Waring | Abbvie, Chicago, IL, United States |
| Nizar Smaoui | Abbvie, Chicago, IL, United States |
| Fedik Rahimov | Abbvie, Chicago, IL, United States |
| Anne Lehtonen | Abbvie, Chicago, IL, United States |
| Keith Usiskin | Celgene, Summit, NJ, United States/ Bristol Myers Squibb, New York, NY, United States |
| Tim Lu | Genentech, San Francisco, CA, United States |
| Natalie Bowers | Genentech, San Francisco, CA, United States |
| Danny Oh | Genentech, San Francisco, CA, United States |
| Sarah Pendergrass | Genentech, San Francisco, CA, United States |
| Linda McCarthy | GlaxoSmithKline, Brentford, United Kingdom |
| Amy Hart | Janssen Research & Development, LLC, Spring House, PA, United States |
| Meijian Guan | Janssen Research & Development, LLC, Spring House, PA, United States |
| Jason Miller | Merck, Kenilworth, NJ, United States |
| Kirsi Kalpala | Pfizer, New York, NY, United States |
| Melissa Miller | Pfizer, New York, NY, United States |
| Xinli Hu | Pfizer, New York, NY, United States |

Large Registry Based Analysis of Genetic Predisposition to Tuberculosis
Identifies Genetic Risk Factors at HLA
Supplements


Rheumatology Group

| | |
|---|---|
| Kari Eklund | Hospital District of Helsinki and Uusimaa, Helsinki, Finland |
| Antti Palomäki | Hospital District of Southwest Finland, Turku, Finland |
| Pia Isomäki | Pirkanmaa Hospital District, Tampere, Finland |
| Laura Pirilä | Hospital District of Southwest Finland, Turku, Finland |
| Oili Kaipiainen-Seppänen | Northern Savo Hospital District, Kuopio, Finland |
| Johanna Huhtakangas | Northern Ostrobothnia Hospital District, Oulu, Finland |
| Nina Mars | Institute for Molecular Medicine Finland, HiLIFE, Helsinki, Finland |
| Ali Abbasi | Abbvie, Chicago, IL, United States |
| Jeffrey Waring | Abbvie, Chicago, IL, United States |
| Fedik Rahimov | Abbvie, Chicago, IL, United States |
| Apinya Lertratanakul | Abbvie, Chicago, IL, United States |
| Nizar Smaoui | Abbvie, Chicago, IL, United States |
| Anne Lehtonen | Abbvie, Chicago, IL, United States |
| David Close | Astra Zeneca, Cambridge, United Kingdom |
| Marla Hochfeld | Celgene, Summit, NJ, United States/ Bristol Myers Squibb, New York, NY, United States |
| Natalie Bowers | Genentech, San Francisco, CA, United States |
| Sarah Pendergrass | Genentech, San Francisco, CA, United States |
| Jorge Esparza Gordillo | GlaxoSmithKline, Brentford, United Kingdom |
| Kirsi Auro | GlaxoSmithKline, Brentford, United Kingdom |
| Dawn Waterworth | Janssen Research & Development, LLC, Spring House, PA, United States |
| Fabiana Farias | Merck, Kenilworth, NJ, United States |
| Kirsi Kalpala | Pfizer, New York, NY, United States |
| Nan Bing | Pfizer, New York, NY, United States |
| Xinli Hu | Pfizer, New York, NY, United States |

Pulmonology Group

| | |
|---|---|
| Tarja Laitinen | Pirkanmaa Hospital District, Tampere, Finland |
| Margit Pelkonen | Northern Savo Hospital District, Kuopio, Finland |

| | |
|---|---|
| Paula Kauppi | Hospital District of Helsinki and Uusimaa, Helsinki, Finland |
| Hannu Kankaanranta | University of Gothenburg, Gothenburg, Sweden/ Seinäjoki Central Hospital, Seinäjoki, Finland/ Tampere University, Tampere, Finland |
| Terttu Harju | Northern Ostrobothnia Hospital District, Oulu, Finland |
| Riitta Lahesmaa | Hospital District of Southwest Finland, Turku, Finland |
| Nizar Smaoui | Abbvie, Chicago, IL, United States |
| Alex Mackay | Astra Zeneca, Cambridge, United Kingdom |
| Glenda Lassi | Astra Zeneca, Cambridge, United Kingdom |
| Susan Eaton | Biogen, Cambridge, MA, United States |
| Steven Greenberg | Celgene, Summit, NJ, United States/ Bristol Myers Squibb, New York, NY, United States |
| Hubert Chen | Genentech, San Francisco, CA, United States |
| Sarah Pendergrass | Genentech, San Francisco, CA, United States |
| Natalie Bowers | Genentech, San Francisco, CA, United States |
| Joanna Betts | GlaxoSmithKline, Brentford, United Kingdom |
| Soumitra Ghosh | GlaxoSmithKline, Brentford, United Kingdom |
| Kirsi Auro | GlaxoSmithKline, Brentford, United Kingdom |
| Rajashree Mishra | GlaxoSmithKline, Brentford, United Kingdom |
| Majd Mouded | Novartis, Basel, Switzerland |
| Debby Ngo | Novartis, Basel, Switzerland |

Cardiometabolic Diseases Group

| | |
|---|---|
| Teemu Niiranen | The National Institute of Health and Welfare Helsinki, Finland |
| Felix Vaura | The National Institute of Health and Welfare Helsinki, Finland |
| Veikko Salomaa | The National Institute of Health and Welfare Helsinki, Finland |
| Kaj Metsärinne | Hospital District of Southwest Finland, Turku, Finland |
| Jenni Aittokallio | Hospital District of Southwest Finland, Turku, Finland |
| Mika Kähönen | Pirkanmaa Hospital District, Tampere, Finland |
| Jussi Hernesniemi | Pirkanmaa Hospital District, Tampere, Finland |
| Juhani Junttila | Northern Ostrobothnia Hospital District, Oulu, Finland |
| Markku Laakso | Northern Savo Hospital District, Kuopio, Finland |

| | |
|---|---|
| Jussi Pihlajamäki | Northern Savo Hospital District, Kuopio, Finland |
| Daniel Gordin | Hospital District of Helsinki and Uusimaa, Helsinki, Finland |
| Juha Sinisalo | Hospital District of Helsinki and Uusimaa, Helsinki, Finland |
| Marja-Riitta Taskinen | Hospital District of Helsinki and Uusimaa, Helsinki, Finland |
| Tiinamaija Tuomi | Hospital District of Helsinki and Uusimaa, Helsinki, Finland |
| Timo Hiltunen | Hospital District of Helsinki and Uusimaa, Helsinki, Finland |
| Jari Laukkanen | Central Finland Health Care District, Jyväskylä, Finland |
| Amanda Elliott | Institute for Molecular Medicine Finland, HiLIFE, University of Helsinki, Finland / Broad Institute, Cambridge, MA, United States |
| Mary Pat Reeve | Institute for Molecular Medicine Finland, HiLIFE, University of Helsinki, Finland |
| Sanni Ruotsalainen | Institute for Molecular Medicine Finland, HiLIFE, University of Helsinki, Finland |
| Benjamin Challis | Astra Zeneca, Cambridge, United Kingdom |
| Dirk Paul | Astra Zeneca, Cambridge, United Kingdom |
| Keith Usiskin | Celgene, Summit, NJ, United States/ Bristol Myers Squibb, New York, NY, United States |
| Julie Hunkapiller | Genentech, San Francisco, CA, United States |
| Natalie Bowers | Genentech, San Francisco, CA, United States |
| Sarah Pendergrass | Genentech, San Francisco, CA, United States |
| Audrey Chu | GlaxoSmithKline, Brentford, United Kingdom |
| Kirsi Auro | GlaxoSmithKline, Brentford, United Kingdom |
| Dermot Reilly | Janssen Research & Development, LLC, Boston, MA, United States |
| Beryl Cummings | Maze Therapeutics, San Francisco, CA, United States |
| Mike Mendelson | Novartis, Boston, MA, United States |
| Jaakko Parkkinen | Pfizer, New York, NY, United States |
| Melissa Miller | Pfizer, New York, NY, United States |

<u>Oncology Group</u>

| | |
|---|---|
| Tuomo Meretoja | Hospital District of Helsinki and Uusimaa, Helsinki, Finland |
| Heikki Joensuu | Hospital District of Helsinki and Uusimaa, Helsinki, Finland |
| Olli Carpén | Hospital District of Helsinki and Uusimaa, Helsinki, Finland |
| Lauri Aaltonen | Hospital District of Helsinki and Uusimaa, Helsinki, Finland |

| | |
|---|---|
| Johanna Mattson | Hospital District of Helsinki and Uusimaa, Helsinki, Finland |
| Eveliina Salminen | Hospital District of Helsinki and Uusimaa, Helsinki, Finland |
| Annika Auranen | Pirkanmaa Hospital District , Tampere, Finland |
| Peeter Karihtala | Northern Ostrobothnia Hospital District, Oulu, Finland |
| Päivi Auvinen | Northern Savo Hospital District, Kuopio, Finland |
| Klaus Elenius | Hospital District of Southwest Finland, Turku, Finland |
| Johanna Schleutker | Hospital District of Southwest Finland, Turku, Finland |
| Esa Pitkänen | Institute for Molecular Medicine Finland, HiLIFE, University of Helsinki, Finland |
| Nina Mars | Institute for Molecular Medicine Finland, HiLIFE, University of Helsinki, Finland |
| Mark Daly | Institute for Molecular Medicine Finland, HiLIFE, University of Helsinki, Finland |
| Relja Popovic | Abbvie, Chicago, IL, United States |
| Jeffrey Waring | Abbvie, Chicago, IL, United States |
| Bridget Riley-Gillis | Abbvie, Chicago, IL, United States |
| Anne Lehtonen | Abbvie, Chicago, IL, United States |
| Jennifer Schutzman | Genentech, San Francisco, CA, United States |
| Julie Hunkapiller | Genentech, San Francisco, CA, United States |
| Natalie Bowers | Genentech, San Francisco, CA, United States |
| Sarah Pendergrass | Genentech, San Francisco, CA, United States |
| Diptee Kulkarni | GlaxoSmithKline, Brentford, United Kingdom |
| Kirsi Auro | GlaxoSmithKline, Brentford, United Kingdom |
| Alessandro Porello | Janssen Research & Development, LLC, Spring House, PA, United States |
| Andrey Loboda | Merck, Kenilworth, NJ, United States |
| Heli Lehtonen | Pfizer, New York, NY, United States |
| Stefan McDonough | Pfizer, New York, NY, United States |
| Marika Crohns | Sanofi, Paris, France |
| Sauli Vuoti | Sanofi, Paris, France |

Opthalmology Group

| | |
|---|---|
| Kai Kaarniranta | Northern Savo Hospital District, Kuopio, Finland |
| Joni A Turunen | Hospital District of Helsinki and Uusimaa, Helsinki, Finland |

| | |
|---|---|
| Terhi Ollila | Hospital District of Helsinki and Uusimaa, Helsinki, Finland |
| Hannu Uusitalo | Pirkanmaa Hospital District, Tampere, Finland |
| Juha Karjalainen | Institute for Molecular Medicine Finland, HiLIFE, University of Helsinki, Finland |
| Esa Pitkänen | Institute for Molecular Medicine Finland, HiLIFE, University of Helsinki, Finland |

| | |
|---|---|
| Mengzhen Liu | Abbvie, Chicago, IL, United States |
| Heiko Runz | Biogen, Cambridge, MA, United States |
| Stephanie Loomis | Biogen, Cambridge, MA, United States |
| Erich Strauss | Genentech, San Francisco, CA, United States |
| Natalie Bowers | Genentech, San Francisco, CA, United States |
| Hao Chen | Genentech, San Francisco, CA, United States |
| Sarah Pendergrass | Genentech, San Francisco, CA, United States |

Dermatology Group

| | |
|---|---|
| Kaisa Tasanen | Northern Ostrobothnia Hospital District, Oulu, Finland |
| Laura Huilaja | Northern Ostrobothnia Hospital District, Oulu, Finland |
| Katariina Hannula-Jouppi | Hospital District of Helsinki and Uusimaa, Helsinki, Finland |
| Teea Salmi | Pirkanmaa Hospital District, Tampere, Finland |
| Sirkku Peltonen | Hospital District of Southwest Finland, Turku, Finland |
| Leena Koulu | Hospital District of Southwest Finland, Turku, Finland |
| Nizar Smaoui | Abbvie, Chicago, IL, United States |
| Fedik Rahimov | Abbvie, Chicago, IL, United States |
| Anne Lehtonen | Abbvie, Chicago, IL, United States |
| David Choy | Genentech, San Francisco, CA, United States |
| Sarah Pendergrass | Genentech, San Francisco, CA, United States |
| Dawn Waterworth | Janssen Research & Development, LLC, Spring House, PA, United States |
| Kirsi Kalpala | Pfizer, New York, NY, United States |
| Ying Wu | Pfizer, New York, NY, United States |

Large Registry Based Analysis of Genetic Predisposition to Tuberculosis
Identifies Genetic Risk Factors at HLA
Supplements

<u>Odontology Group</u>


| | |
|---|---|
| Pirkko Pussinen | Hospital District of Helsinki and Uusimaa, Helsinki, Finland |
| Aino Salminen | Hospital District of Helsinki and Uusimaa, Helsinki, Finland |
| Tuula Salo | Hospital District of Helsinki and Uusimaa, Helsinki, Finland |
| David Rice | Hospital District of Helsinki and Uusimaa, Helsinki, Finland |
| Pekka Nieminen | Hospital District of Helsinki and Uusimaa, Helsinki, Finland |
| Ulla Palotie | Hospital District of Helsinki and Uusimaa, Helsinki, Finland |
| Maria Siponen | Northern Savo Hospital District, Kuopio, Finland |
| Liisa Suominen | Northern Savo Hospital District, Kuopio, Finland |
| Päivi Mäntylä | Northern Savo Hospital District, Kuopio, Finland |
| Ulvi Gursoy | Hospital District of Southwest Finland, Turku, Finland |
| Vuokko Anttonen | Northern Ostrobothnia Hospital District, Oulu, Finland |
| Kirsi Sipilä | Northern Ostrobothnia Hospital District, Oulu, Finland |
| Sarah Pendergrass | Genentech, San Francisco, CA, United States |


<u>Women's Health and Reproduction Group</u>


| | |
|---|---|
| Hannele Laivuori | Institute for Molecular Medicine Finland, HiLIFE, University of Helsinki, Finland |
| Venla Kurra | Pirkanmaa Hospital District, Tampere, Finland |
| Laura Kotaniemi-Talonen | Pirkanmaa Hospital District, Tampere, Finland |
| Oskari Heikinheimo | Hospital District of Helsinki and Uusimaa, Helsinki, Finland |
| Ilkka Kalliala | Hospital District of Helsinki and Uusimaa, Helsinki, Finland |
| Lauri Aaltonen | Hospital District of Helsinki and Uusimaa, Helsinki, Finland |
| Varpu Jokimaa | Hospital District of Southwest Finland, Turku, Finland |
| Johannes Kettunen | Northern Ostrobothnia Hospital District, Oulu, Finland |
| Marja Vääräsmäki | Northern Ostrobothnia Hospital District, Oulu, Finland |
| Outi Uimari | Northern Ostrobothnia Hospital District, Oulu, Finland |
| Laure Morin-Papunen | Northern Ostrobothnia Hospital District, Oulu, Finland |
| Maarit Niinimäki | Northern Ostrobothnia Hospital District, Oulu, Finland |
| Terhi Piltonen | Northern Ostrobothnia Hospital District, Oulu, Finland |
| Katja Kivinen | Institute for Molecular Medicine Finland, HiLIFE, University of Helsinki, Finland |

| | |
|---|---|
| Elisabeth Widen | Institute for Molecular Medicine Finland, HiLIFE, University of Helsinki, Finland |
| Taru Tukiainen | Institute for Molecular Medicine Finland, HiLIFE, University of Helsinki, Finland |
| Mary Pat Reeve | Institute for Molecular Medicine Finland, HiLIFE, University of Helsinki, Finland |
| Mark Daly | Institute for Molecular Medicine Finland, HiLIFE, University of Helsinki, Finland |
| Liu Aoxing | Institute for Molecular Medicine Finland, HiLIFE, University of Helsinki, Finland |
| Andrea Ganna | Institute for Molecular Medicine Finland, HiLIFE, University of Helsinki, Finland |
| Niko Välimäki | University of Helsinki, Helsinki, Finland |
| Eija Laakkonen | University of Jyväskylä, Jyväskylä, Finland |
| Jaakko Tyrmi | University of Oulu, Oulu, Finland / University of Tampere, Tampere, Finland |
| Heidi Silven | University of Oulu, Oulu, Finland |
| Eeva Slitz | University of Oulu, Oulu, Finland |
| Riikka Arffman | University of Oulu, Oulu, Finland |
| Susanna Savukoski | University of Oulu, Oulu, Finland |
| Triin Laisk | Estonian biobank, Tartu, Estonia |
| Natalia Pujol | Estonian biobank, Tartu, Estonia |
| Bridget Riley-Gillis | Abbvie, Chicago, IL, United States |
| Mengzhen Liu | Abbvie, Chicago, IL, United States |
| Sarah Pendergrass | Genentech, San Francisco, CA, United States |
| Janet Kumar | GlaxoSmithKline, Brentford, United Kingdom |
| Kirsi Auro | GlaxoSmithKline, Brentford, United Kingdom |


#### FinnGen Analysis working group


| | |
|---|---|
| Bridget Riley-Gillis | Abbvie, Chicago, IL, United States |
| Reza Hammond | Abbvie, Chicago, IL, United States |
| Fedik Rahimov | Abbvie, Chicago, IL, United States |
| Sabah Kadri | Abbvie, Chicago, IL, United States |
| Mengzhen Liu | Abbvie, Chicago, IL, United States |
| Slavé Petrovski | Astra Zeneca, Cambridge, United Kingdom |
| Eleonor Wigmore | Astra Zeneca, Cambridge, United Kingdom |

Large Registry Based Analysis of Genetic Predisposition to Tuberculosis
Identifies Genetic Risk Factors at HLA
Supplements


| | |
|---|---|
| Adele Mitchell | Biogen, Cambridge, MA, United States |
| Benjamin Sun | Biogen, Cambridge, MA, United States |
| Ellen Tsai | Biogen, Cambridge, MA, United States |
| Denis Baird | Biogen, Cambridge, MA, United States |
| Paola Bronson | Biogen, Cambridge, MA, United States |
| Ruoyu Tian | Biogen, Cambridge, MA, United States |
| Stephanie Loomis | Biogen, Cambridge, MA, United States |
| Yunfeng Huang | Biogen, Cambridge, MA, United States |
| Till Andlauer | Boehringer Ingelheim, Ingelheim am Rhein, Germany |
| Jatin Arora | Boehringer Ingelheim, Ingelheim am Rhein, Germany |
| Ghadi Rai | Boehringer Ingelheim, Ingelheim am Rhein, Germany |
| Zhihao Ding | Boehringer Ingelheim, Ingelheim am Rhein, Germany |
| Lorenz Maier | Boehringer Ingelheim, Ingelheim am Rhein, Germany |
| Karsten Quast | Boehringer Ingelheim, Ingelheim am Rhein, Germany |
| Francisco Herruzo | Boehringer Ingelheim, Ingelheim am Rhein, Germany |
| Daniel Lopez | Boehringer Ingelheim, Ingelheim am Rhein, Germany |
| Marc Jung | Boehringer Ingelheim, Ingelheim am Rhein, Germany |
| Boris Bartholdy | Boehringer Ingelheim, Ingelheim am Rhein, Germany |
| Joseph Maranville | Celgene, Summit, NJ, United States/ Bristol Myers Squibb, New York, NY, United States |
| Shameek Biswas | Celgene, Summit, NJ, United States/ Bristol Myers Squibb, New York, NY, United States |
| Elmutaz Mohammed | Celgene, Summit, NJ, United States/ Bristol Myers Squibb, New York, NY, United States |
| Samir Wadhawan | Celgene, Summit, NJ, United States/ Bristol Myers Squibb, New York, NY, United States |
| Erika Kvikstad | Celgene, Summit, NJ, United States/ Bristol Myers Squibb, New York, NY, United States |
| Minal Caliskan | Celgene, Summit, NJ, United States/ Bristol Myers Squibb, New York, NY, United States |
| Diana Chang | Genentech, San Francisco, CA, United States |
| Julie Hunkapiller | Genentech, San Francisco, CA, United States |

Large Registry Based Analysis of Genetic Predisposition to Tuberculosis
Identifies Genetic Risk Factors at HLA
Supplements


| | |
|---|---|
| Tushar Bhangale | Genentech, San Francisco, CA, United States |
| Natalie Bowers | Genentech, San Francisco, CA, United States |
| Sarah Pendergrass | Genentech, San Francisco, CA, United States |
| Karen S King | GlaxoSmithKline, Brentford, United Kingdom |
| Padhraig Gormley | GlaxoSmithKline, Brentford, United Kingdom |
| Jimmy Liu | GlaxoSmithKline, Brentford, United Kingdom |
| Karsten Sieber | Janssen Research & Development, LLC, Spring House, PA, United States |
| Amy Hart | Janssen Research & Development, LLC, Spring House, PA, United States |
| Meijian Guan | Janssen Research & Development, LLC, Spring House, PA, United States |
| Shicheng Guo | Janssen Research & Development, LLC, Spring House, PA, United States |
| Beryl Cummings | Maze Therapeutics, San Francisco, CA, United States |
| Matt Brauer | Maze Therapeutics, San Francisco, CA, United States |
| Jason Miller | Merck, Kenilworth, NJ, United States |
| Fabiana Farias | Merck, Kenilworth, NJ, United States |
| Jorge Del-Aguila | Merck, Kenilworth, NJ, United States |
| Kirill Shkura | Merck, Kenilworth, NJ, United States |
| Victor Neduva | Merck, Kenilworth, NJ, United States |
| Huilei Xu | Novartis, Basel, Switzerland |
| Amy Cole | Novartis, Basel, Switzerland |
| Jonathan Chung | Novartis, Basel, Switzerland |
| Jaison Jacob | Novartis, Basel, Switzerland |
| Katrina de Lange | Novartis, Basel, Switzerland |
| Jonas Zierer | Novartis, Basel, Switzerland |
| Xing Chen | Pfizer, New York, NY, United States |
| Åsa Hedman | Pfizer, New York, NY, United States |
| Clarence Wang | Sanofi, Paris, France |
| Ethan Xu | Sanofi, Paris, France |
| Franck Auge | Sanofi, Paris, France |
| Clement Chatelain | Sanofi, Paris, France |
| Deepak Rajpal | Sanofi, Paris, France |

Large Registry Based Analysis of Genetic Predisposition to Tuberculosis
Identifies Genetic Risk Factors at HLA
Supplements


| | |
|---|---|
| Dongyu Liu | Sanofi, Paris, France |
| Katherine Call | Sanofi, Paris, France |
| Tai-He Xia | Sanofi, Paris, France |
| Mitja Kurki | Institute for Molecular Medicine Finland, HiLIFE, University of Helsinki, Finland / Broad Institute, Cambridge, MA, United States |
| Samuli Ripatti | Institute for Molecular Medicine Finland, HiLIFE, University of Helsinki, Finland |
| Mark Daly | Institute for Molecular Medicine Finland, HiLIFE, University of Helsinki, Finland |
| Juha Karjalainen | Institute for Molecular Medicine Finland, HiLIFE, University of Helsinki, Finland |
| Aki Havulinna | Institute for Molecular Medicine Finland, HiLIFE, University of Helsinki, Finland |
| Juha Mehtonen | Institute for Molecular Medicine Finland, HiLIFE, University of Helsinki, Finland |
| Priit Palta | Institute for Molecular Medicine Finland, HiLIFE, University of Helsinki, Finland |
| Shabbeer Hassan | Institute for Molecular Medicine Finland, HiLIFE, University of Helsinki, Finland |
| Pietro Della Briotta Parolo | Institute for Molecular Medicine Finland, HiLIFE, University of Helsinki, Finland |
| Wei Zhou | Broad Institute, Cambridge, MA, United States |
| Mutaamba Maasha | Broad Institute, Cambridge, MA, United States |
| Shabbeer Hassan | Institute for Molecular Medicine Finland, HiLIFE, University of Helsinki, Finland |
| Susanna Lemmelä | Institute for Molecular Medicine Finland, HiLIFE, University of Helsinki, Finland |
| Manuel Rivas | University of Stanford, Stanford, CA, United States |
| Aarno Palotie | Institute for Molecular Medicine Finland, HiLIFE, University of Helsinki, Finland |
| Arto Lehisto | Institute for Molecular Medicine Finland, HiLIFE, University of Helsinki, Finland |
| Andrea Ganna | Institute for Molecular Medicine Finland, HiLIFE, University of Helsinki, Finland |
| Vincent Llorens | Institute for Molecular Medicine Finland, HiLIFE, University of Helsinki, Finland |
| Hannele Laivuori | Institute for Molecular Medicine Finland, HiLIFE, University of Helsinki, Finland |
| Taru Tukiainen | Institute for Molecular Medicine Finland, HiLIFE, University of Helsinki, Finland |
| Mary Pat Reeve | Institute for Molecular Medicine Finland, HiLIFE, University of Helsinki, Finland |
| Henrike Heyne | Institute for Molecular Medicine Finland, HiLIFE, University of Helsinki, Finland |
| Nina Mars | Institute for Molecular Medicine Finland, HiLIFE, University of Helsinki, Finland |
| Kimmo Palin | University of Helsinki, Helsinki, Finland |
| Javier Garcia-Tabuenca | University of Tampere, Tampere, Finland |
| Harri Siirtola | University of Tampere, Tampere, Finland |
| Tuomo Kiiskinen | Institute for Molecular Medicine Finland, HiLIFE, University of Helsinki, Finland |

Large Registry Based Analysis of Genetic Predisposition to Tuberculosis
Identifies Genetic Risk Factors at HLA
Supplements


| | |
|---|---|
| Jiwoo Lee | Institute for Molecular Medicine Finland, HiLIFE, University of Helsinki, Finland / Broad Institute, Cambridge, MA, United States |
| Kristin Tsuo | Institute for Molecular Medicine Finland, HiLIFE, University of Helsinki, Finland / Broad Institute, Cambridge, MA, United States |
| Amanda Elliott | Institute for Molecular Medicine Finland, HiLIFE, University of Helsinki, Finland / Broad Institute, Cambridge, MA, United States |
| Kati Kristiansson | THL Biobank / The National Institute of Health and Welfare Helsinki, Finland |
| Mikko Arvas | Finnish Red Cross Blood Service / Finnish Hematology Registry and Clinical Biobank, Helsinki, Finland |
| Kati Hyvärinen | Finnish Red Cross Blood Service, Helsinki, Finland |
| Jarmo Ritari | Finnish Red Cross Blood Service, Helsinki, Finland |
| Miika Koskinen | Helsinki Biobank / Helsinki University and Hospital District of Helsinki and Uusimaa, Helsinki |
| Olli Carpén | Helsinki Biobank / Helsinki University and Hospital District of Helsinki and Uusimaa, Helsinki |
| Johannes Kettunen | Northern Finland Biobank Borealis / University of Oulu / Northern Ostrobothnia Hospital District, Oulu, Finland |
| Katri Pylkäs | University of Oulu, Oulu, Finland |
| Eeva Sliz | University of Oulu, Oulu, Finland |
| Minna Karjalainen | University of Oulu, Oulu, Finland |
| Tuomo Mantere | Northern Finland Biobank Borealis / University of Oulu / Northern Ostrobothnia Hospital District, Oulu, Finland |
| Eeva Kangasniemi | Finnish Clinical Biobank Tampere / University of Tampere / Pirkanmaa Hospital District, Tampere, Finland |
| Sami Heikkinen | University of Eastern Finland, Kuopio, Finland |
| Arto Mannermaa | Biobank of Eastern Finland / University of Eastern Finland / Northern Savo Hospital District, Kuopio, Finland |
| Eija Laakkonen | University of Jyväskylä, Jyväskylä, Finland |
| Dhanaprakash Jambulingam | University of Turku, Turku, Finland |
| Venkat Subramaniam Rathinakannan | University of Turku, Turku, Finland |
| Nina Pitkänen | Auria Biobank / University of Turku / Hospital District of Southwest Finland, Turku, Finland |

Large Registry Based Analysis of Genetic Predisposition to Tuberculosis
Identifies Genetic Risk Factors at HLA
Supplements

#### Biobank directors


| | |
|---|---|
| Lila Kallio | Auria Biobank / University of Turku / Hospital District of Southwest Finland, Turku, Finland |
| Sirpa Soini | THL Biobank / The National Institute of Health and Welfare Helsinki, Finland |
| Jukka Partanen | Finnish Red Cross Blood Service / Finnish Hematology Registry and Clinical Biobank, Helsinki, Finland |
| Eero Punkka | Helsinki Biobank / Helsinki University and Hospital District of Helsinki and Uusimaa, Helsinki |
| Raisa Serpi | Northern Finland Biobank Borealis / University of Oulu / Northern Ostrobothnia Hospital District, Oulu, Finland |
| Sanna Siltanen | Finnish Clinical Biobank Tampere / University of Tampere / Pirkanmaa Hospital District, Tampere, Finland |
| Veli-Matti Kosma | Biobank of Eastern Finland / University of Eastern Finland / Northern Savo Hospital District, Kuopio, Finland |
| Teijo Kuopio | Central Finland Biobank / University of Jyväskylä / Central Finland Health Care District, Jyväskylä, Finland |


#### FinnGen Teams

Administration


| | |
|---|---|
| Anu Jalanko | Institute for Molecular Medicine Finland, HiLIFE, University of Helsinki, Finland |
| Huei-Yi Shen | Institute for Molecular Medicine Finland, HiLIFE, University of Helsinki, Finland |
| Risto Kajanne | Institute for Molecular Medicine Finland, HiLIFE, University of Helsinki, Finland |
| Mervi Aavikko | Institute for Molecular Medicine Finland, HiLIFE, University of Helsinki, Finland |


Analysis


| | |
|---|---|
| Mitja Kurki | Institute for Molecular Medicine Finland, HiLIFE, University of Helsinki, Finland / Broad Institute, Cambridge, MA, United States |
| Juha Karjalainen | Institute for Molecular Medicine Finland, HiLIFE, University of Helsinki, Finland |
| Pietro Della Briotta Parolo | Institute for Molecular Medicine Finland, HiLIFE, University of Helsinki, Finland |
| Arto Lehisto | Institute for Molecular Medicine Finland, HiLIFE, University of Helsinki, Finland |
| Juha Mehtonen | Institute for Molecular Medicine Finland, HiLIFE, University of Helsinki, Finland |

| | |
|---|---|
| Wei Zhou | Broad Institute, Cambridge, MA, United States |
| Masahiro Kanai | Broad Institute, Cambridge, MA, United States |
| Mutaamba Maasha | Broad Institute, Cambridge, MA, United States |

Clinical Endpoint Development

| | |
|---|---|
| Hannele Laivuori | Institute for Molecular Medicine Finland, HiLIFE, University of Helsinki, Finland |
| Aki Havulinna | Institute for Molecular Medicine Finland, HiLIFE, University of Helsinki, Finland |
| Susanna Lemmelä | Institute for Molecular Medicine Finland, HiLIFE, University of Helsinki, Finland |
| Tuomo Kiiskinen | Institute for Molecular Medicine Finland, HiLIFE, University of Helsinki, Finland |
| L. Elisa Lahtela | Institute for Molecular Medicine Finland, HiLIFE, University of Helsinki, Finland |

Communication

| | |
|---|---|
| Mari Kaunisto | Institute for Molecular Medicine Finland, HiLIFE, University of Helsinki, Finland |

E-Science

| | |
|---|---|
| Elina Kilpeläinen | Institute for Molecular Medicine Finland, HiLIFE, University of Helsinki, Finland |
| Timo P. Sipilä | Institute for Molecular Medicine Finland, HiLIFE, University of Helsinki, Finland |
| Oluwaseun Alexander Dada | Institute for Molecular Medicine Finland, HiLIFE, University of Helsinki, Finland |
| Awaisa Ghazal | Institute for Molecular Medicine Finland, HiLIFE, University of Helsinki, Finland |
| Anastasia Shcherban | Institute for Molecular Medicine Finland, HiLIFE, University of Helsinki, Finland |
| Rigbe Weldatsadik | Institute for Molecular Medicine Finland, HiLIFE, University of Helsinki, Finland |

Genotyping

| | |
|---|---|
| Kati Donner | Institute for Molecular Medicine Finland, HiLIFE, University of Helsinki, Finland |
| Timo P. Sipilä | Institute for Molecular Medicine Finland, HiLIFE, University of Helsinki, Finland |

Sample Collection Coordination

| | |
|---|---|
| Anu Loukola | Helsinki Biobank / Helsinki University and Hospital District of Helsinki and Uusimaa, Helsinki |

Large Registry Based Analysis of Genetic Predisposition to Tuberculosis
Identifies Genetic Risk Factors at HLA
Supplements

Sample Logistics


| | |
|---|---|
| Päivi Laiho | THL Biobank / The National Institute of Health and Welfare Helsinki, Finland |
| Tuuli Sistonen | THL Biobank / The National Institute of Health and Welfare Helsinki, Finland |
| Essi Kaiharju | THL Biobank / The National Institute of Health and Welfare Helsinki, Finland |
| Markku Laukkanen | THL Biobank / The National Institute of Health and Welfare Helsinki, Finland |
| Elina Järvensivu | THL Biobank / The National Institute of Health and Welfare Helsinki, Finland |
| Sini Lähteenmäki | THL Biobank / The National Institute of Health and Welfare Helsinki, Finland |
| Lotta Männikkö | THL Biobank / The National Institute of Health and Welfare Helsinki, Finland |
| Regis Wong | THL Biobank / The National Institute of Health and Welfare Helsinki, Finland |


Registry Data Operations


| | |
|---|---|
| Hannele Mattsson | THL Biobank / The National Institute of Health and Welfare Helsinki, Finland |
| Kati Kristiansson | THL Biobank / The National Institute of Health and Welfare Helsinki, Finland |
| Susanna Lemmelä | Institute for Molecular Medicine Finland, HiLIFE, University of Helsinki, Finland |
| Sami Koskelainen | THL Biobank / The National Institute of Health and Welfare Helsinki, Finland |
| Tero Hiekkalinna | THL Biobank / The National Institute of Health and Welfare Helsinki, Finland |
| Teemu Paajanen | THL Biobank / The National Institute of Health and Welfare Helsinki, Finland |


Sequencing Informatics


| | |
|---|---|
| Priit Palta | Institute for Molecular Medicine Finland, HiLIFE, University of Helsinki, Finland |
| Kalle Pärn | Institute for Molecular Medicine Finland, HiLIFE, University of Helsinki, Finland |
| Mart Kals | Institute for Molecular Medicine Finland, HiLIFE, University of Helsinki, Finland |
| Shuang Luo | Institute for Molecular Medicine Finland, HiLIFE, University of Helsinki, Finland |
| Vishal Sinha | Institute for Molecular Medicine Finland, HiLIFE, University of Helsinki, Finland |


Trajectory


| | |
|---|---|
| Tarja Laitinen | Pirkanmaa Hospital District, Tampere, Finland |
| Mary Pat Reeve | Institute for Molecular Medicine Finland, HiLIFE, University of Helsinki, Finland |
| Marianna Niemi | University of Tampere, Tampere, Finland |
| Harri Siirtola | University of Tampere, Tampere, Finland |

Large Registry Based Analysis of Genetic Predisposition to Tuberculosis
Identifies Genetic Risk Factors at HLA
Supplements

Javier Gracia-Tabuenca University of Tampere, Tampere, Finland

Mika Helminen University of Tampere, Tampere, Finland

Tiina Luukkaala University of Tampere, Tampere, Finland

Iida Vähätalo University of Tampere, Tampere, Finland

Data protection officer

Jyrki Pitkänen Institute for Molecular Medicine Finland, HiLIFE, University of Helsinki, Finland

**FINBB - Finnish biobank cooperative**

Marco Hautalahti

Johanna Mäkelä

Mirkka Koivusalo

Sarah Smith

Tom Southerington
